# Gene expression signatures identify pediatric patients with multiple organ dysfunction who require advanced life support in the intensive care unit

**DOI:** 10.1101/2020.02.15.20022772

**Authors:** Rama Shankar, Mara L. Leimanis, Patrick A. Newbury, Ke Liu, Jing Xing, Derek Nedveck, Eric J. Kort, Jeremy W Prokop, Guoli Zhou, André S Bachmann, Bin Chen, Surender Rajasekaran

## Abstract

**Background:** Multiple organ dysfunction syndrome (MODS) occurs in the setting of a variety of pathologies including infection and trauma. Some of these patients will further decompensate and require extra corporeal membrane oxygenation (ECMO) as a palliating maneuver to allow time for recovery of cardiopulmonary function. The molecular mechanisms driving progression from MODS to cardiopulmonary collapse remain incompletely understood, and no biomarkers have been defined to identify those MODS patients at highest risk for progression to requiring ECMO support. We hypothesize that molecular features derived from whole blood transcriptomic profiling either alone or in combination with traditional clinical and laboratory markers can prospectively identify these high risk MODS patients in the pediatric intensive care unit (PICU).

**Design/Methods:** Whole blood RNA-seq profiling was performed for 23 MODS patients at three time points during their ICU stay (at diagnosis of MODS, 72 hours after, and 8 days later), as well as four healthy controls undergoing routine sedation. Of the 23 MODS patients, six required ECMO support (ECMO patients). The predictive power of conventional demographic and clinical features was quantified for differentiating the MODS and ECMO patients. We then compared the performance of markers derived from transcriptomic profiling including (1) transcriptomically imputed leukocyte subtype distribution, (2) relevant published gene signatures and (3) a novel differential gene expression signature computed from our data set. The predictive power of our novel gene expression signature was then validated using independently published datasets.

**Results:** None of the five demographic characteristics and 14 clinical features, including The Pediatric Logistic Organ Dysfunction (PELOD) score, could predict deterioration of MODS to ECMO at baseline. From previously published sepsis signatures, only the signatures positively associated with patients mortality could differentiate ECMO patients from MODS patients, when applied to our transcriptomic dataset (*P*-value ranges from 0.01 to 0.04). Deconvolution of bulk RNA-Seq samples suggested that lower neutrophil counts were associated with increased risk of progression from MODS to ECMO (*P*-value = 0.03, OR=2.82 [95% CI 0.63– 12.45]). A total of 28 genes were differentially expressed between ECMO and MODS patients at baseline (log_2_ fold change ≥ 1 or ≤ -1 with false discovery rate ≤ 0.2). These genes are involved in protein maintenance and epigenetic-related processes. Further univariate analysis of these 28 genes suggested a signature of six DE genes associated with ECMO (OR > 3.0, *P*-value ≤ 0.05). Notably, this contains a set of histone marker genes, including *H1F0, HIST2H3C, HIST1H2AI, HIST1H4*, and *HIST1H1B*, that were highly expressed in ECMO. A risk score derived from expression of these genes differentiated ECMO and MODS patients in our dataset (AUC = 0.91, 95% CI 0.79-0.1.00, *P*-value = 7e-04) as well as validation dataset (AUC= 0.73,95% CI 0.53-0.93, *P*-value = 2e-02).

**Conclusions:** This study identified lower neutrophils and upregulation of specific histone related genes as a putative signature for deterioration of MODS to ECMO. This study demonstrates that transcriptomic features may be superior to traditional clinical methods of ascertaining severity in patients with MODS.

**Author summary:** *Why was this study done?:*

- Multiple organ dysfunction syndrome (MODS) is a major cause of mortality and morbidity in critically ill pediatric patients who survive the initial physical insult.
- A variety of triggers including trauma and infections can lead to MODS in pediatric patients.
- The clinical condition of some MODS patients improve while others deteriorate, needing resource-intensive life support such as extracorporeal membrane oxygenation (ECMO).
- Mortality is uncommon in PICUs and the need for advanced life support devices, such as ECMO can serve as proxy for mortality.
- The decision to initiate ECMO in pediatric patients is often subjective made by a committee of physicians that include surgeons, intensivists and a variety of other subspecialists often in the absence of objective data.
- Despite decades of research, no diagnostic criteria or biomarker has been identified that comprehensively assesses severity in MODS patients who may need subsequent ECMO support in the hyperacute phase of injury.
- We systematically assessed clinical and transcriptional features as biomarkers for the prediction of the ECMO patients.

*What did the researcher do and find?:*

- We investigated various clinical and transcriptional features in 27 patients with MODS at multiple time points (4 CT, 17 MODS, 6 ECMO) at baseline (0h).
- We observed that immune response pathways (monocytes, cytokines, NF-kB, and inflammation) were activated in the initiation of MODS, whereas neutrophil level was decreased during deterioration of MODS to ECMO.
- A total of 51 DE genes were identified in MODS compared to CT and 28 DE in ECMO compared to MODS at baseline (0h).
- We identified the enrichment of immune-related and glycogenolysis processes in MODS compared to CT and enrichment of protein maintenance, DNA repair and epigenetic-related processes in ECMO compared to MODS at baseline (0h).
- Logistic regression was used to identify a signature of 6 genes strongly associated with ECMO decision and this signature could help to diagnose MODS patients requiring ECMO.
- The transcriptomic signature-based risk scores were further evaluated in an independent cohort.

*What do these findings mean?:*

- The compromised level of neutrophils and activation of gene markers including a few histone genes could be used as putative signature for diagnosing the deterioration of MODS to ECMO.
- A risk score derived from signature genes could be used to predict the need for ECMO.
- This score is superior to traditional clinical criteria and severity scores used in the Pediatric ICU.
- The transcriptional signature derived in this study could potentially be used to identify patients in the hyperacute phase of injury that may need higher levels of support like ECMO enabling the selection of an appropriate treatment plan.

## Introduction

Multiple organ dysfunction syndrome (MODS) is common in the pediatric intensive care unit (PICU), being diagnosed in the majority of patients with sepsis as well as many trauma patients [1]. MODS complicates a wide range of pathologies including severe hypoxemia, cardiorespiratory arrest, shock, trauma, acute pancreatitis, gut malperfusion, acute leukemia, solid organ or hematopoietic stem cell transplantation, hemophagocytic lymphohistiocytosis, and thrombotic microangiopathy [2]. Contemporary management of MODS is entirely supportive, and focused on addressing the underlying disease process.

Some pediatric patients who develop MODS deteriorate and require intensive life support in the form of Extracorporeal Membrane Oxygenation (ECMO). It has been observed that pediatrics patients requiring ECMO have a 50-60% mortality rate [3]. The decision to initiate ECMO remains subjective based on the empirical experience of the multidisciplinary care team. Establishment of objective markers of what patients will require ECMO support would simplify the decision making process and potentially enable earlier intervention for these patients. However, no clinical scoring tool or molecular biomarker has been developed to identify the patients who may require subsequent advanced support; therefore, developing biomarkers for identifying MODS patients at high risk of requiring ECMO support is a critical unmet need.

Whole blood transcriptomic profiling has been evaluated to perform risk-stratification of sepsis patients, predict mortality in sepsis and better understand the pathogenesis of MODS [4]. A number of published gene expression signatures shed light on the molecular mechanism of MODS [5,6]. However, none of the signatures were developed with a view towards identifying patients that require ECMO support.

In this work, we present a cohort of MODS patients, a subset of whom progressed to requiring ECMO support (MODS vs ECMO) and healthy controls (CT). Here we use the term MODS to denote those MODS patients that did not require ECMO and ECMO for MODS patients deteriorated to needing ECMO support. These patients were assessed using a combination of conventional demographic and clinical markers, as well as whole blood transcriptomic profiling in an effort to identify diagnostic markers that can distinguish between the MODS and ECMO patient population.

## Methods

### Patients and blood sampling

The IRB of this study (2016-062-SH/HDVCH) was approved by Spectrum Health on May 17, 2016. All the patients were minors and their parents were consented prior to recruitment into our study. Every parent gave consent. After IRB approval, a short-term longitudinal design was adopted to assess the transcriptomic profiles of patients from the PICU at Helen DeVos Children’s Hospital, Michigan. Critically ill patients, meeting criteria for MODS as determined by clinical observations, were screened for eligibility and consented. Blood samples were collected at three time points: at recognition of MODS (0h), 72 hours after, and 8 days later (N=27). Samples were collected in PaxGene® tubes and stored at -80°C. Healthy controls (N=4) were patients that presented for same day sedation. Samples from each control patient were obtained only once and were reported as 0h. Of the 23 MODS patients, 6 required ECMO support. From admission to day 8, 47% of the MODS patients were discharged to home or out of the ICU to a medical floor. Patients who left the ICU did not have further blood draws.One patient from the ECMO group died during the study and two other MODS patients died six months later.

### Sequencing

RNA samples were prepared using KAPA RNA HyperPrep Kit, and sequenced on an Illumina NextSeq500. Using ribosomal reduction RNAseq methodology, we were able to capture both cellular and acellular RNA signatures of all PICU patients.

### Validation Data Sets

For validation, we were unable to identify any analogous publicly available gene expression datasets that included pediatric MODS patients at multiple time points. We therefore chose a dataset describing an adult cohort (23-63 years) developed MODS in the hyperacute phase of trauma [7]. This dataset was used as an independent cohort to validate our signature genes. The MODS patients in this validation dataset were categorized into MODS and noMODS (patients did not develop MODS) as described in patient demographics [7]. In addition, a single cell RNA-Seq dataset was also available for adult ECMO patients [8]. We used the immune cell markers from this dataset to validate our immune response analysis.

## Bioinformatics analysis

### RNA-Seq data analysis

All the sequencing reads were mapped on Hg38 transcriptome using the ENSEMBL GRCh38.p3 annotation with the STAR aligner [9]. The edgeR package [10] was used for quantification of differentially expressed (DE) genes with criteria: log_2_ fold change ≥ 1 or ≤ -1 with adjusted *P*-value (False Discover Rate) < 0.20. DE genes were identified between the two groups in all the three-time points separately. The DE genes were used for co-expression network analysis using CEMiTools package [11]. The gene ontology (GO) enrichment of DE genes was performed using the clusterProfiler R package [12]. Biological processes with adjusted *P*-value ≤ 0.01 were considered as significantly enriched. Dotplot function provided in clusterProfiler was used to visualize enriched pathways. In addition, gene interaction network was visualized using STRING: functional protein association network (https://string-db.org/).

### Immune Cell Deconvolution

CIBERSORT was used to estimate the relative composition of immune cells in bulk RNA-Seq samples [13] using a machine learning model named as nu–support vector regression (*v*-SVR) [14]. For each patient, a complete blood count (CBC) was obtained upon presentation as part of their standard of care clinical evaluation. We were therefore able to calculate estimated absolute counts for each leukocyte subpopulation. This was done by multiplying the proportion for each subpopulation as determined by CIBERSORT to the total white blood cell count from the CBC. This analysis was validated by comparing the absolute neutrophil counts (ANC) as estimated by CIBERSORT with the ANC reported by the clinical laboratory.

### Statistical Analysis

All plots and statistical analyses were carried out using R programming language (v3.5.1) (https://www.r-project.org/). By default, two-sided student’s t-test was performed % to compute the significance between two groups. The generalized linear model function (glm) was used to calculate odds ratio (OR). Principal component analysis (PCA) of gene expression profiles was performed using the prcomp function. The risk score was estimated using the signature gene expression for each patient based on the geometric mean. The geometric mean for *x*_1_, *x*_2_, …, *x_n_* was calculated as follows:

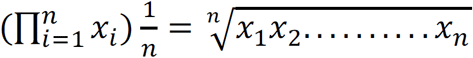

A risk score was further used to re-classify patients into two groups and receiver operating characteristic (ROC) and area under curve (AUC) were adopted to assess the performance using the pROC package [15].

## Results

The workflow of the study is summarized in Fig. 1. Patient demographics and baseline clinical parameters are provided in Table 1, with 72h and 8 day values presented in supplemental Table S1. In total, five demographic characteristics (i.e., age, gender, BMI, weight, height) and 14 different clinical features were examined for all patients. There was high variation between MODS and ECMO for many clinical parameters (e.g. platelet count), diluting the predictive power of these measures. Some outcomes differed significantly between MODS and ECMO, specifically the renal failure rate (89% in MODS and 100% in ECMO) and liver failure rate (30% in MODS and 50% in ECMO. However, no baseline demographic or clinical parameter, including PELOD score was predictive of progression from MODS to ECMO. This observation highlighted the need to explore molecular features for identifying risk markers.

**Table 1:** Patients demographics at baseline (Pre-ECMO,0h) time point.

| RNA-Seq cohort |  |  |  |  |
| --- | --- | --- | --- | --- |
| Demographics | Control | MODS | ECMO | P-value |
| Time | 0h | 0h | 0h | - |
| Number | 4 | 17 | 6 | - |
| Age (months) | 84.75(28-122) | 90(0.14-202) | 63.25(0.5-202) | 0.54 |
| Male | 2 | 10 | 5 | 0.36 |
| Female | 2 | 7 | 1 | 0.36 |
| BMI | 17(14-21) | 20.3(13-38.5) | 19(14-32.4) | 0.74 |
| Weight | 26.5(12-35) | 42.85(3.5-178) | 25.87(3.9-81) | 0.35 |
| Height | 122(80-142) | 103(51-157) | 90(53-160) | 0.59 |
| Mortality | - | 2 | 1 |  |
| Clinical Features |  |  |  |  |
| Liver Failure (%) | - | 30 | 50 | - |
| Bilirubin | - | 0.92(0.1-5.6) | 0.51(0.1-1.1) | 0.28 |
| AST | - | 258.88(13-3296) | 215.67(7-726) | 0.85 |
| Albumin | - | 2.35(1.6-3.5) | 2.35(1.9-2.8) | 0.96 |
| CRP | - | 83.75(0.3-234) | 75.75(2.8-211) | 0.87 |
| Renal Failure (%) | - | 89 | 100 | - |
| Creatinine | - | 0.65(0.13-0.29) | 0.77(0.22-0.29) | 0.71 |
| Lactate | - | 2.2(0.9-4.6) | 6.05(0.6-14.5) | 0.19 |
| WBC | - | 14.9(3.95-62.6) | 12.67(4.6-20) | 0.56 |
| platelet | - | 232(37-718) | 208(92-378) | 0.68 |
| PELOD Score | - | 14.37(1-32) | 12.5(10-20) | 0.21 |
| Bacterial infection (%) | - | 35 | 33 |  |
| Viral infection (%) | - | 52 | 50 |  |
| Inotrope usage |  | 88 | 100 |  |
| Respiratory failure (%) | - | 100 | 100 |  |
| Neurological (%) | - | 23 | 33 |  |

Where relevant, mean(range). T-test and fisher's exact test was used to compute *P*-value between MODS and ECMO.

**Fig. 1.**
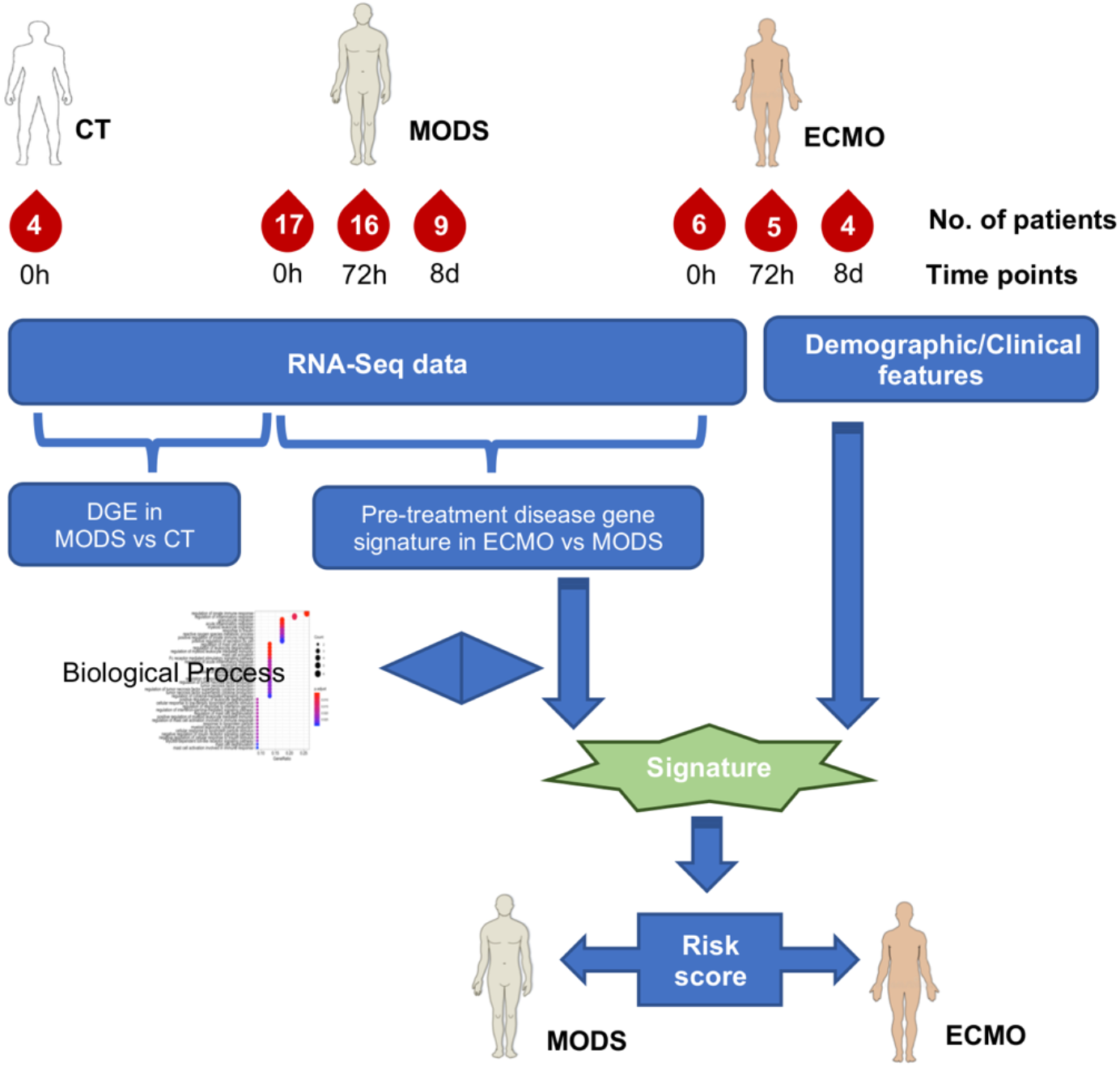
An overview of the analysis. DGE: Differential gene expression.

### Immune cells deconvolution and transcriptome analysis

Immune responses were examined for individual patients and compared to elucidate their role. The relative proportions of immune cell subtypes were estimated using CIBERSORT based on bulk RNA-Seq data. WBC counts obtained upon arrival in the emergency department were used to quantify the absolute abundance of immune cell subtypes. The ANC as determined by the clinical laboratory and the ANC derived from CIBERSORT were high correlated (correlation value 0.97) (Figure S1), suggesting the high fidelity of the inferred leukocyte subtype composition. Comparison of neutrophils between ECMO and MODS showed decreased level in ECMO (*P*-value = 0.03, OR=2.82 [95% CI 0.63 – 12.45]) as compared to MODS (Fig. 2a). Interestingly, the two lowest neutrophil counts were among MODS. Clinical data of these two patients revealed that one patient did not survive and another had the PELOD score of 32, the highest score among all patients, suggesting that these patients had a risk profile similar to the ECMO patients despite not being started on ECMO.

**Fig. 2.**
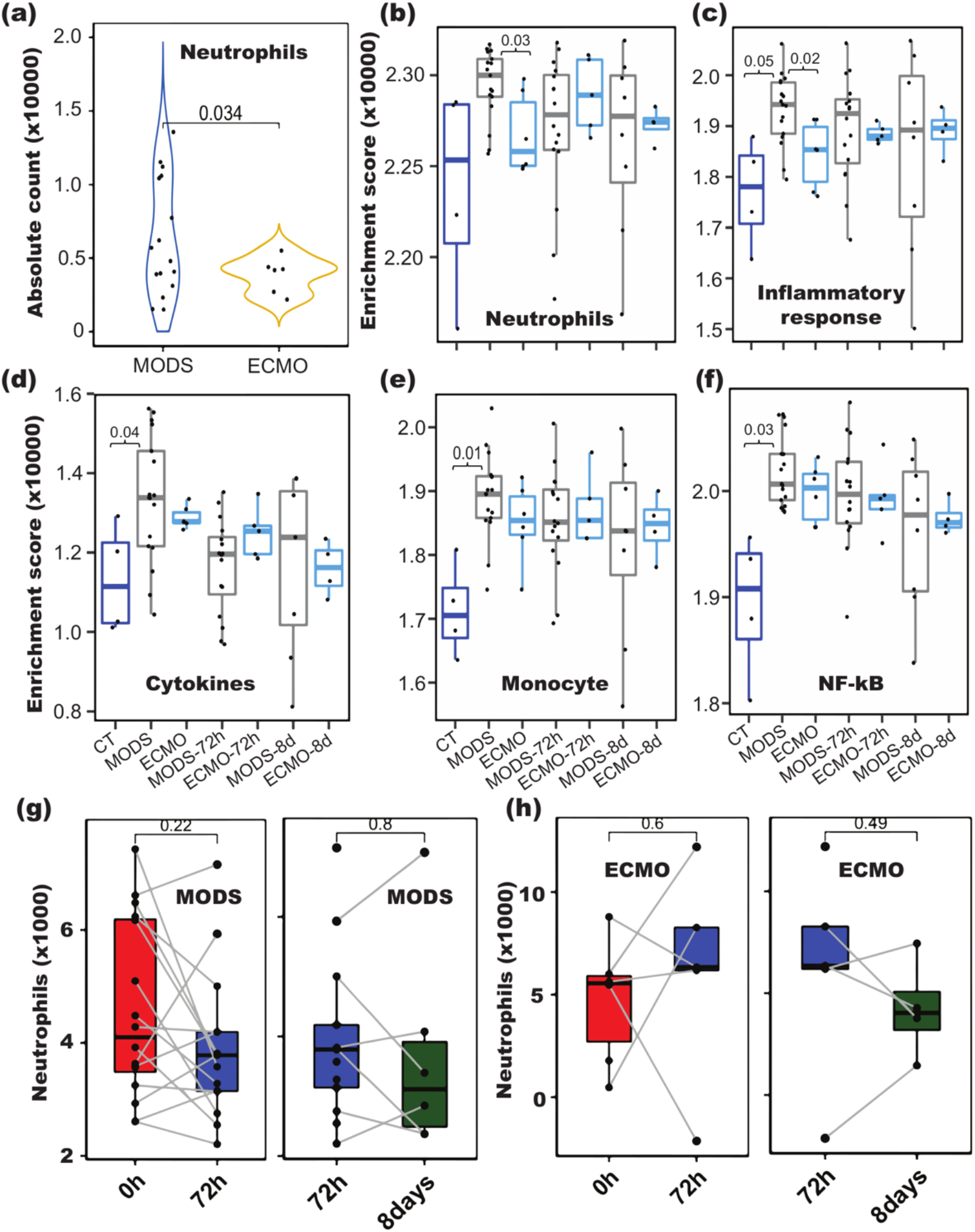
Immune cell composition analyses in ECMO and MODS patients. (a) Neutrophils counts computed from CIBERSORT decreased in ECMO (P = 0.034) compared to MODS at baseline. (b-f) Enrichment of genes involved in various immune responses (Monocytes, Cytokines, NF-kB, Neutrophils and Inflammation) in CT, MODS and ECMO at different time points (0h, 72h and 8d). Abundance of neutrophils in MODS (g) and ECMO (h) patients at different time points (0h, 72h and 8days). Blue color - control (CT), grey color - MODS patients and cyan color - ECMO patients.

We then examined the expression of marker genes of neutrophils (from CIBERSORT), monocytes, cytokines and genes involved in NF-kB and inflammatory response from Hall et al., 2007 [16]. All the marker genes were down-regulated in ECMO compared to MODS (Figure S2-S6). In addition to CIBERSORT, gene set enrichment analysis of cell-type specific biomarker genes was performed in order to confirm the findings. Neutrophil gene markers and genes involved in inflammatory response displayed significantly decreased expression in ECMO compared to MODS (*P*-value < 0.03) (Fig. 2b and 2c). Marker genes pertaining to monocytes, cytokines, and NF-kB displayed a significant enrichment in MODS compared to CT (*P*-value < 0.04) (Fig. 2d-2f).

The finding of changes in the neutrophil count was independently validated using additional single cell RNA-seq data of ECMO adult patients data [8], where we observed decrease of expression of neutrophil gene markers and genes involved in inflammatory response in deceased ECMO patients compared to patients that survived (Figure S7 and S8). Further paired comparison of neutrophil levels for each patient showed no significant change across different time points (Fig. 2g and 2h).

Furthermore, differential expression (DE) analysis between MODS and control (CT) as well as between ECMO and MODS was performed at baseline (0h). A total of 51 DE genes (log_2_ fold change ≥ 1 or ≤ -1 with false discovery rate (FDR) ≤ 0.2) between MODS and CT, and 28 DE genes between ECMO and MODS were identified at baseline (Fig. 3a). Comparison of DE genes from these two groups showed only one pseudogene (RNU1-67P) common to these two DE lists. As expected, these DE genes clearly separate CT, MODS and ECMO patients (Fig. 3b and 3c) in reduced dimensional (PC) space. Heatmap visualization of these genes highlights their differential patterns of expression between groups (Fig. 3d).

**Fig. 3.**
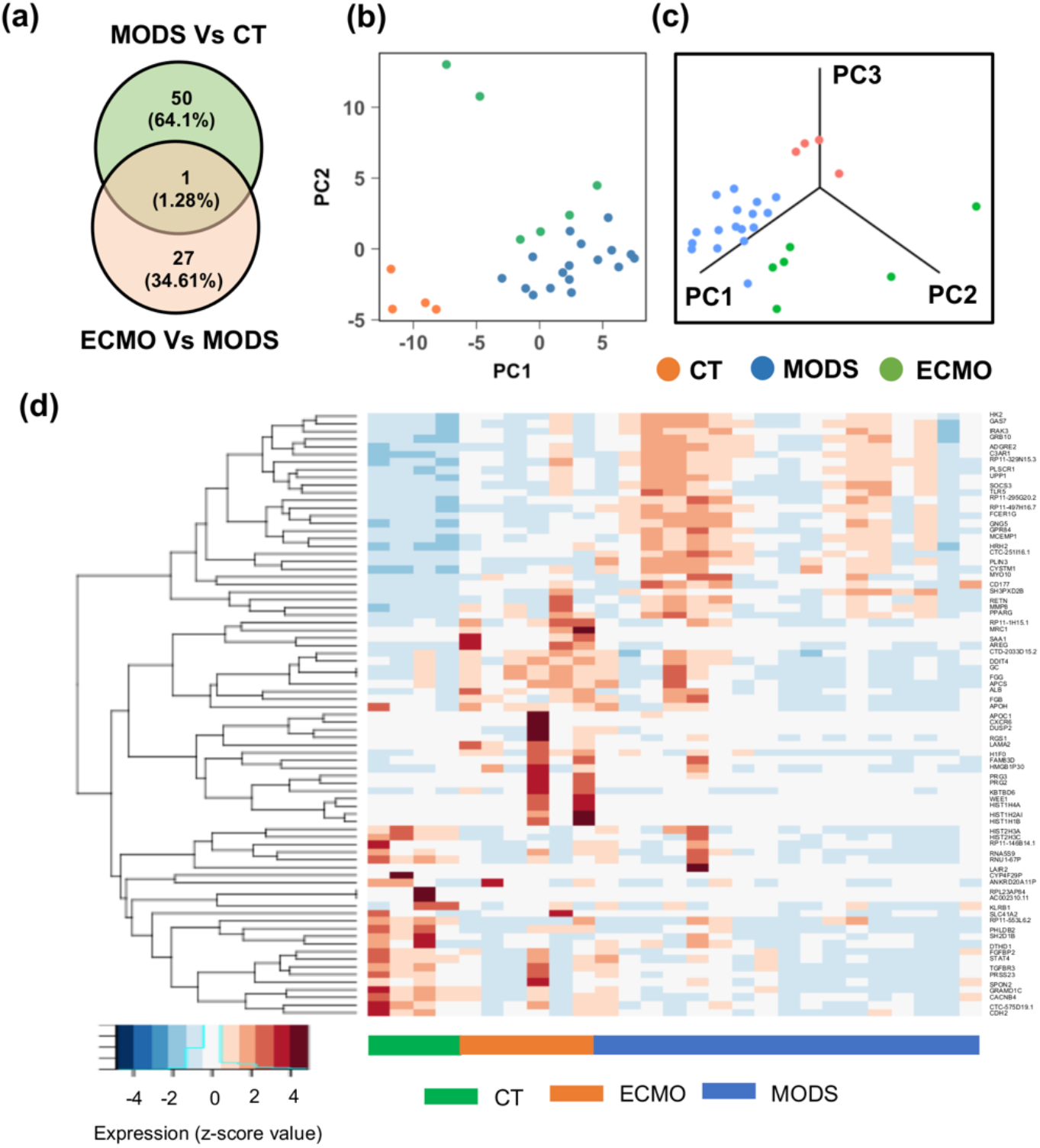
Differential gene expression analyses at baseline (0h). (a) Comparison of differentially expressed (DE) genes between MODS vs. control (CT), and ECMO vs. MODS at baseline (0h). (b) First two principal components and (c) first three principal component analysis, using the union of DE genes obtained from the comparison between MODS and CT and that those between ECMO and MODS at baseline. Patients are clustered by their pathology group (CT, MODS and ECMO). (d) Expression of the DE genes.

In addition, 50 and 32 DE genes between MODS and CT were identified at 72h and 8d time points, respectively (Figure S9a), while 9 and 11 DE genes were identified between ECMO and MODS at 72h and 8d time points, respectively (Figure S9b). Only one gene (pseudogene-RNU1-67P) was common among all the three time points (0h, 72h and 8d) in both comparisons.

## Biological processes and co-expression networks regulated by DE genes

Gene ontology (GO) enrichment analysis of total DE genes from MODS to CT comparison revealed that immune-related (innate immune response, mast cell activation, neutrophil migration, interleukin-6 production, and cytokine production) and fatty acid-related pathways are enriched in MODS compared to CT (corrected *P*-value ≤ 0.01, Fig. 4a) (Table S3). Notable genes included in immune responses are *ADGRE2, C3AR1, CD177, FCER1G, IRAK3, MMP8, PLSCR1, PPARG, SOCS3*, and *TLR5*. Similar pathways were also observed in a previous analysis between MODS and CT [5]. In addition, gene expression related to epigenetic processes (e.g., regulation of gene silence, DNA packaging, chromatin assembly) was activated in ECMO compared to MODS (Fig. 4b and Table S4).

**Fig. 4.**
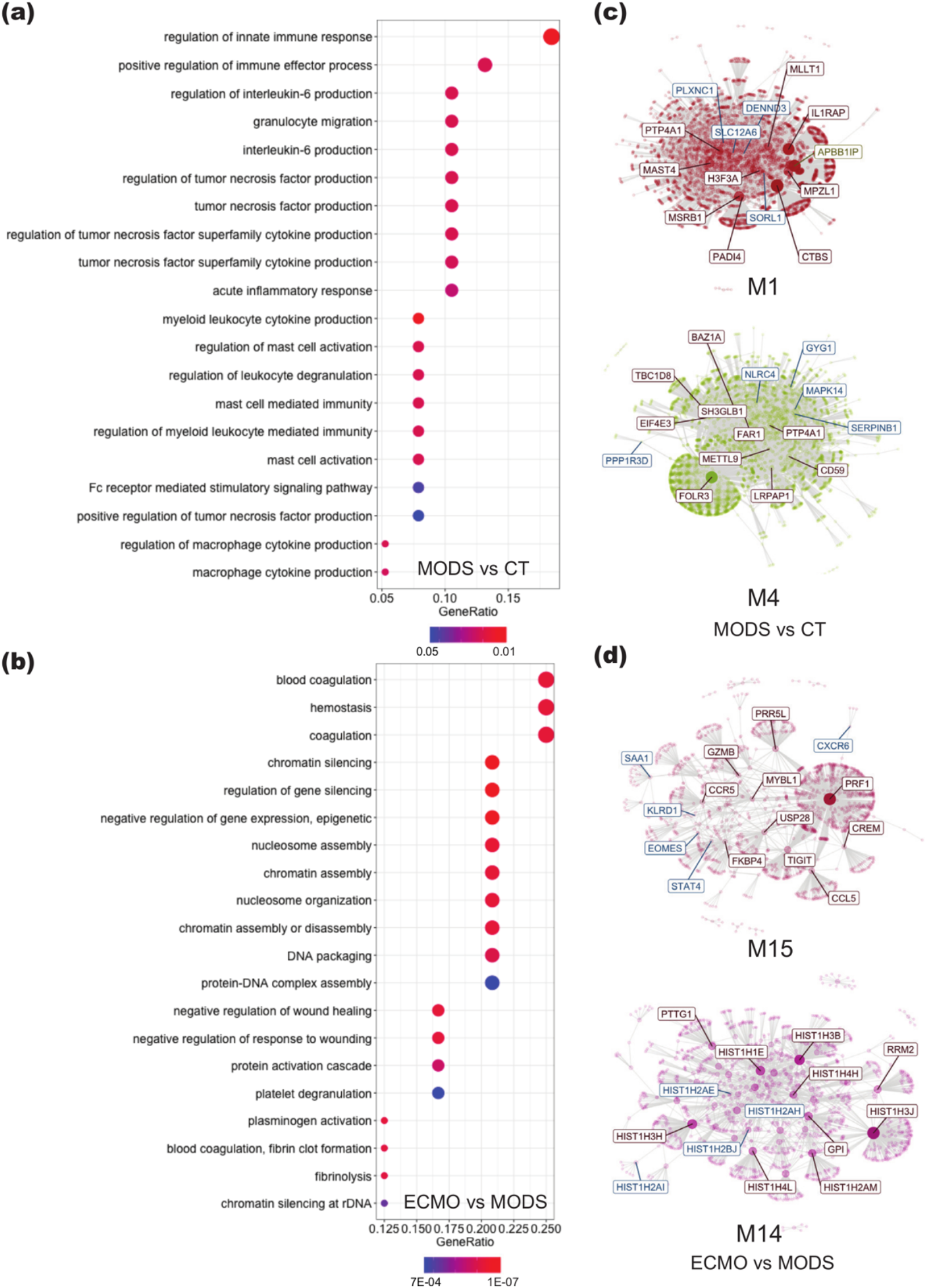
Gene enrichment and co-expression network analysis of DE genes in MODS and CT, and in ECMO and MODS. (a) Gene ontology (GO) enrichment of DE genes from MODS to CT showed their involvement in immune responses. (b) However, GO enrichment of DE genes from ECMO to MODS displayed enrichment in epigenetic regulations. (c) The DE genes obtained from the comparison of MODS and CT were clustered into two separate groups. (d) Similarly, two co-expression networks were created after mapping the DE genes in ECMO and MODS. The highlighted genes in co-expressed networks are hub genes. Notably, many DE genes from both comparisons were shared in module 15 (M15), suggested phase transition. Size of circles in GO represents the number of mapped genes.

Further, co-expression analysis was performed to delineate the relationships between gene expression and their regulated pathways. The TPM count of all the genes for baseline patients was used to create co-expressed network modules. The DE genes from MODS to CT and ECMO to MODS were mapped on these modules and identified the corresponding modules. Two modules were identified in each comparison (Fig.4c and 4d). Notably, some of the DE genes from MODS to CT were mapped on module M15 of ECMO to MODS, deciphering the phase transition of MODS to ECMO support. Module M14 was specific to the comparison of ECMO and MODS, whereas modules M1 and M4 were specific to the comparison of MODS and CT. Pathways analysis of each module showed that genes in module M1 were involved in immune responses (Figure S10a) and genes in module M4 were involved in glucose metabolisms and glycogen breakdown (Figure S10b). However, module M15 (shared by both comparisons) showed enrichment of signaling pathways and proteins maintenance (Figure S10c). Module M14 belonging to genes that differed between ECMO and MODS was enriched with genes related to DNA damage, DNA maintenance and histone acetylation (Figure S10d). Together, DE analysis showed enrichment of immune related and glycogenolysis pathways in MODS, while protein maintenance and epigenetic-related pathways were enriched in ECMO. The protein-protein interaction network of the DE genes also revealed two distinct clusters: histone activation and blood coagulation were uniquely enriched in ECMO (Figure S11).

The GO enrichment analysis and co-expression analysis of DE genes expressed at 72h and 8d did not show any significantly enriched pathway in any of the comparisons. This observation may suggest that the MODS and ECMO patients have important physiological differences at baseline, but that other processes obfuscate these differences as diverse disease processes and therapeutic interventions unfold. Such baseline differences could be exploited for prognostic and potentially diagnostic purposes.

### Identification of molecular signatures associated with ECMO

In 2018, Sweeney et al. [4] evaluated four prognostic biomarker signatures consisting of genes positively or negatively correlated with mortality in sepsis. We computed the geometric mean of the expression of these signature genes and investigated whether these values could be used as risk scores for MODS to ECMO progression. We observed that the risk scores derived from the signature genes that are positively correlated with mortality among sepsis patients could differentiate ECMO and MODS (*P*-value ranges from 0.04 to 0.01) (Figure S12).

We next sought to derived the predictive power of the differentially expressed genes identified between ECMO and MODS. Six genes from our differential gene expression analysis demonstrated a very strong association with MODS for their progression to ECMO (*P-*value < 0.04, Fig. 5a) and these were used to create a signature for ECMO prediction. Most of these genes belong to the histone family (*HIST2H3C, HIST1H4A, HIST1H2AI, HIST1H1B, and H1F0*, Table 2) and these were expressed significantly higher in ECMO than MODS (P-value < 3.5e-6, Fig. 5b). In addition, the Human Protein Atlas dataset showed the enhanced expression of these genes in neutrophils (Figure S14).

**Table 2:** List of signature genes strongly associated with ECMO.

| Gene | Ensemble | Log<sub>2</sub><br>Fold<br>change | Log<br>CPM | P-value | Adj. P-value | Protein<br>coding | Function |
| --- | --- | --- | --- | --- | --- | --- | --- |
| HIST2H3C | ENSG00000203811 | 2.11 | 3.88 | 9.10E-07 | 0.055 | Y | histone cluster 2, H3c |
| HIST1H4A | ENSG00000278637 | 1.85 | 0.24 | 1.00E-06 | 0.061 | Y | histone cluster 1, H4a |
| HIST1H2AI | ENSG00000196747 | 1.62 | 3.86 | 3.40E-06 | 0.207 | Y | histone cluster 1, H2ai |
| HIST1H1B | ENSG00000184357 | 1.92 | 4.24 | 2.40E-06 | 0.147 | Y | histone cluster 1, H1b |
| H1F0 | ENSG00000189060 | 1.73 | 4.5 | 3.50E-06 | 0.212 | Y | H1 histone family, member 0 |
| DDIT4 | ENSG00000168209 | 2.02 | 3.74 | 4.40E-07 | 0.026 | Y | DNA-damage-inducible transcript 4 |

**Fig. 5.**
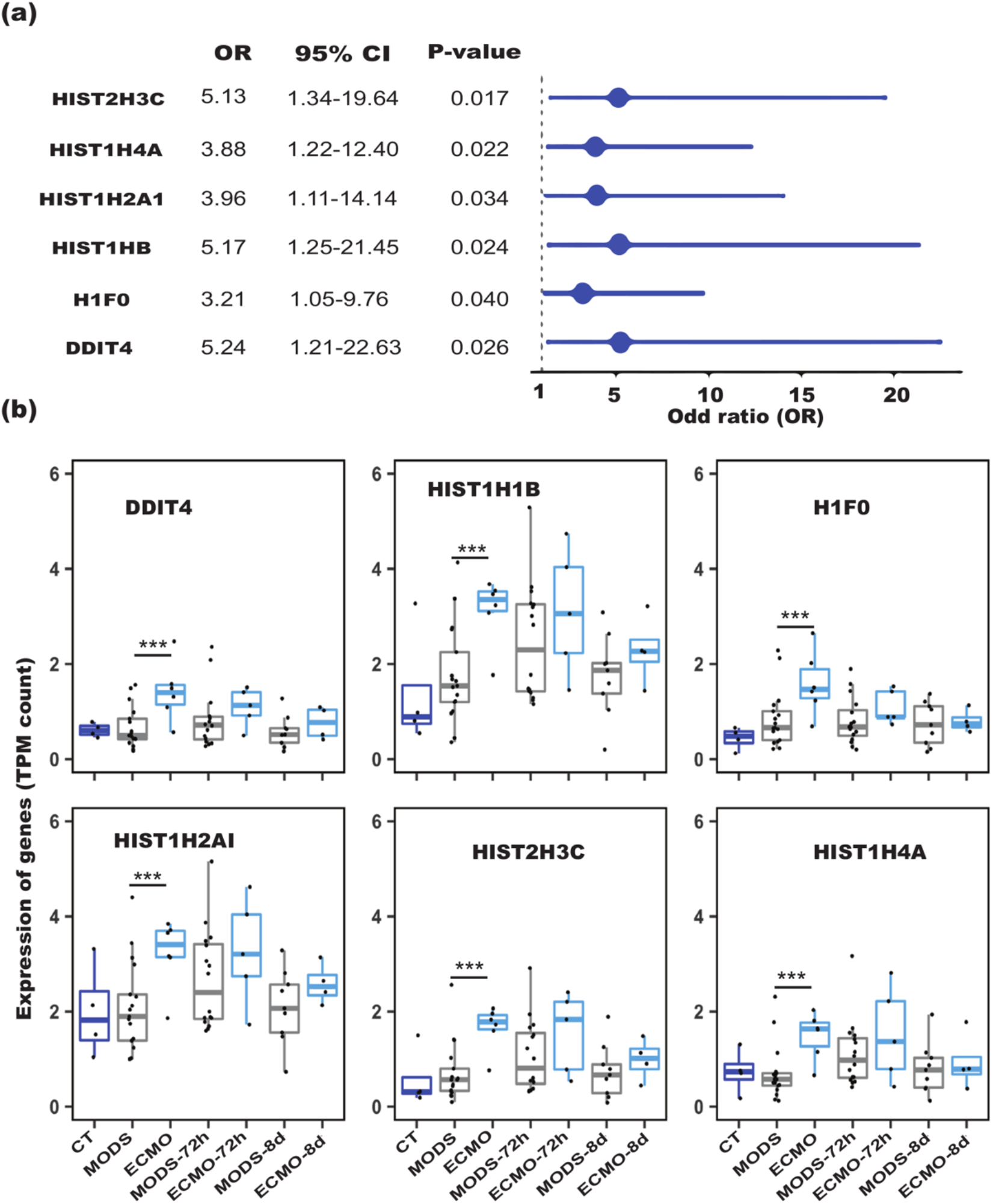
Univariate analyses of differentially expressed (DE) genes in ECMO and MODS. (a) Odds ratio of the DE genes between ECMO and MODS (reference). A total of 6 genes from 28 DE genes are significant (OR > 1 and P value < 0.05). (b) Expression of the DE genes in CT, ECMO and MODS patients at different time points. The higher expression of the genes in ECMO than in MODS at three time points (0h, 72h and 8d) suggests their strong association with the deterioration from MODS to ECMO. Blue color displayed-control (CT), grey color displayed- MODS patients and cyan color displayed- ECMO patients.*** P-value < 1E-06.

### Re-classification of patients and signature-based risk estimation

Expression of the genes in our 6 gene risk signature was similar between CT and MODS, but higher in ECMO than MODS (Fig. 6a). Interestingly, when the additional time points (72h and 8d) were added, these signature genes were not different in MODS and ECMO and could also be confirmed by the overlap of patients (Figure S15a and S15b). The risk scores derived from these genes were significantly different between ECMO and MODS (95%CI 1.54-42.91,*P-*value = 7E-04, Fig. 6b) at baseline. In contrast, risk scores of MODS patients at 72h and 8d are close to those of ECMO patients at 72h and 8d (Fig. 6b).

**Fig. 6.**
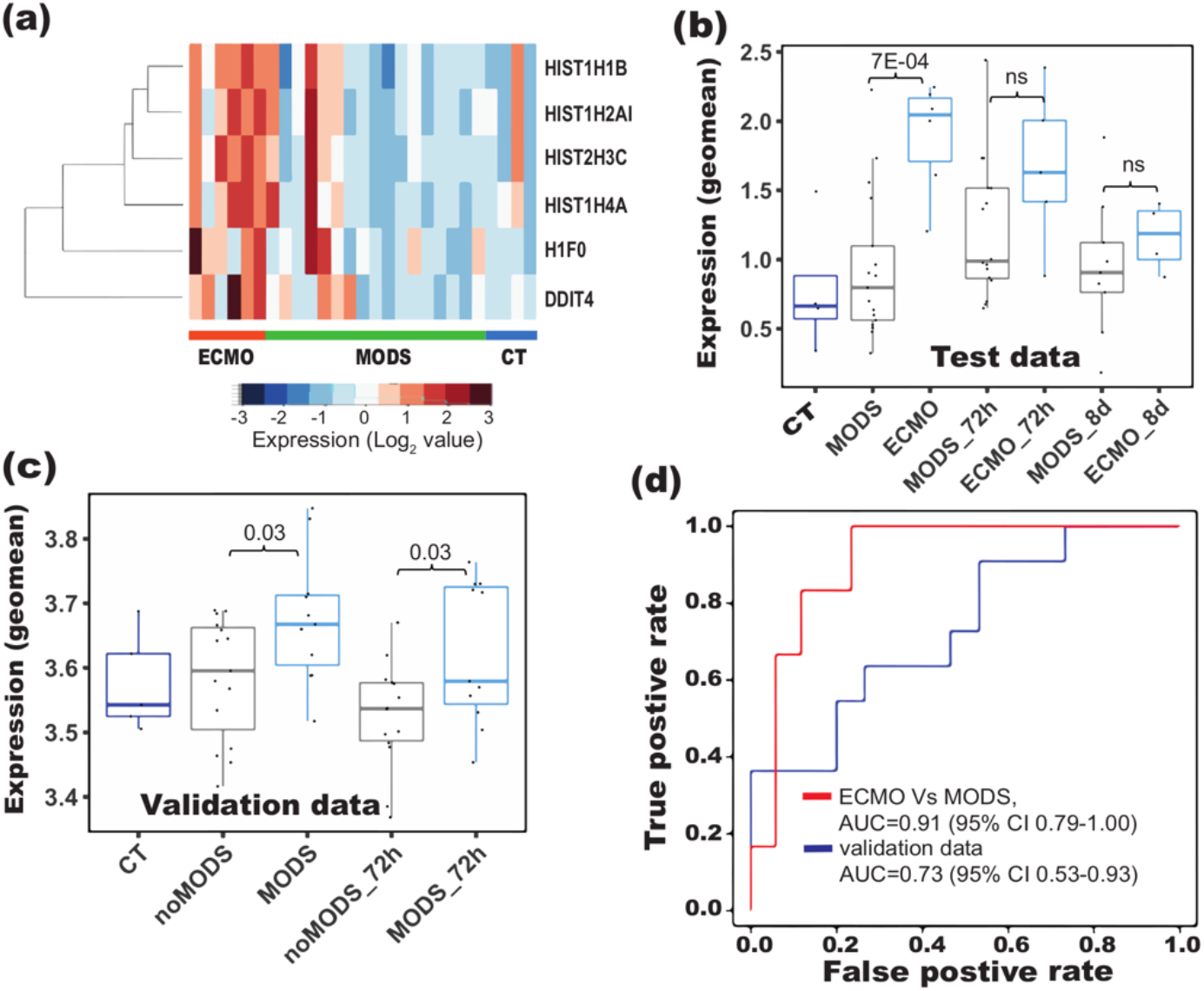
Signature based re-classification of patients in the test (CT, MODS and ECMO) dataset and validation dataset. (a) Heatmaps showed the clustering of signature genes in ECMO patients compared to control (CT) and MODS patients. Risk scores derived from the signature genes showed difference in (b) ECMO and MODS in our data, and in (c) MODS and noMODS (patients doesn’t develop MODS) in the validation data (Cabrera et at., 2017). (d) Receiver operating characteristics (ROC) of the classification using our data and the validation data. A risk score for each patient was computed based on the geometric mean of the signature gene expression. Risk scores were strongly associated with ECMO and can be helpful to predict the probability of the MODS patients who require ECMO support.

Due to the lack of an appropriate pediatric cohort, we used previously published microarray data of adult patients that developed MODS after a major trauma as validation data. The authors had categorized the patients into two groups, those that developed MODS and those that did not (noMODS), however these were more sick compared to controls [9]. In their cohort, the risk score derived from our signature was significantly higher (95%CI 1.02-10.35,*P-*value = 2E-02) in MODS than noMODS (Fig. 6c). We further found that our signature genes can also classify patients (noMODS, and MODS) in the validation cohort at 0h (Figure S17a) as well as 72h timepoint (Figure S17b). Using logistic regression to train the risk scores led to a remarkable separation (AUC of 0.91 [95%CI 0.79-1.00] for ECMO and MODS patients at baseline in our data and AUC of 0.8 [95%CI 0.53-0.93] in the validation set) of two group of patients from our data as well as validation data, indicating a strong association of risk scores with MODS deterioration (Fig. 6d).

## Discussion

The decision to initiate ECMO is often subjective, determined by the clinical judgement of the multidisciplinary care team in a very stressful and dynamic setting as opposed to quantitative measures of pathophysiology. Biological sampling of the exact organs affected is impractical if not impossible, but circulating white blood cells may serve as a proxy read out of stressed be experienced by multiple organ systems. We employed transcriptomics of peripheral white cells in an effort to improve our understanding of the response of circulating cells to multi-organ failure and its progression to either recovery or cardiopulmonary collapse culminating in the need for extra corporeal life support.

White blood cells are uniquely suited for this because aside from a few exceptions (e.g., memory T cells, some tissue macrophages), most of the mature blood cell types are mitotically inactive, metabolically active and relatively short-lived with half-lives ranging over hours to a few days. Thus, they are reflective of the environment they course through [17]. We found the gene *AREG* which regulates Amphiregulin a mediator for macrophage activity were preferentially activated in patients prior to ECMO [18-19]. Amphiregulin has been shown to an essential cardioprotective mediator produced by cardiac Ly6C macrophages in response to fluid overload, which is not unusual in MODS [20].

The activation of immune response and glycogenolysis in MODS compared to CT showed that patients in MODS need excessive energy for cellular homeostasis and activation of immune response against the initial infections. However, during the transition from MODS to ECMO, various signaling and protein maintenance pathways also got activated. Notably, DNA repair, DNA methylation and other epigenetic changes were activated in the patients who deceased further and needed ECMO support.

One of the key observations is the enrichment and strong association of histone genes with ECMO. The histone octamer *HIST2H3C, HIST1H2AI, HIST1H4*, and *HIST1H1B*, are genes that increase the availability of histones. Among these histones, *HIST2H3C, HIST1H2AI*, and *HIST1H4A* are highly expressed in neutrophils (Figure S14). Histones are a protein class, containing histone H1 and the core histones H2A, H2B, H3, and H4 [21] that are involved in numerous biological processes, largely through repressing transcription [22-23]. These are important due to their capability to determine if DNA is accessible for transcription and they have a major impact on gene expression, too [24]. However, to allow processes like transcription or replication, this structure needs to change dynamically from a condensed state to an open one.

Genes that are associated with the histone cluster were found to be elevated. Increases in serum histones have previously been shown to be elevated in patients with sepsis and heart failure [25-26]. Higher concentrations of circulatory histones are associated with poor survival in patients undergoing ECMO [27]. The increased availability of histones in pathologies that concur with a prolonged inflammatory response as is the case of sepsis. This is not only due to tissue damage but also to a second source: activated neutrophils generate neutrophil extracellular traps (NETs), structures made of cellular components which include specifically modified histones [28]. Generation of circulating histones from NETs or from necrotic neutrophils implies the release of a high concentration of histones to the bloodstream. Both processes, NET and apoptosis of neutrophils and necrosis of neutrophils and other immune cells, contribute to the pathogenesis of sepsis. NET however has been linked to organ failure [29-31]. In this study we showed that these processes are active enough to be uncovered by gene-expression.

This study shows that serial whole-blood transcriptomic profiling holds a great promise to predict MODS patients, which may need EMCO support. Several published gene signatures developed to predict mortality showed a significance in predicting ECMO, but none of them suffice as a marker in our case. Our new signature genes could remarkably differentiate MODS and ECMO. Their association with ECMO is considerably strong and is also able to distinguish the severe and moderate MODS patients in the validation cohort. The risk score derived from the signature genes for each patient can be used to classify patients into two groups (ECMO and MODS) in our cohort. This is important because in spite of the limited sample size, using pediatric ECMO samples, the multiple time points and validation datasets increase the robustness of our findings. Furthermore the study included patients, where sepsis was not the primary cause of MODS indicating that histone signatures that occur in patients with MODS do so regardless of the initial insult. The signature genes need further evaluation by prospective studies in pediatric MODS/ECMO patients. Nevertheless, this study is one of the first to demonstrate that the potential of exploring clinical and transcriptomic features in identifying MODS patients from those requiring ECMO. In addition, this work may be of some help to guide the treatment of those infected patients at highest risk for progression to requiring ECMO support.

## Data and Code Availability

The codes used in this analyses are available at https://github.com/Bin-Chen-Lab/MODS. The processed data used in this study is available through NCBI GEO accession GSE144406.

## Acknowledgement

Research reported in this publication was supported by the National Institute Of General Medical Sciences of the National Institutes of Health under Award Number R01GM134307. The content is solely the responsibility of the authors and does not necessarily represent the official views of the National Institutes of Health. We would like to thank Dr. Hui Shen for critical comments and the Van Andel Genomics Core for providing sequencing facilities and services.

## Author contributions

Conceived and designed the experiments: BC, SR and RS. Performed the experiments: RS, ML and DM,. Analyzed the data: RS and PN. Contributed material/analysis tools: KL, JX, EK, JWP, GZ, ASB. Wrote the paper: RS, BC, EK, SR and ML. Supervised the study: BC and SR.

## Abbreviation

MODS: Multiple organ dysfunction syndrome
ECMO: Extracorporeal Membrane Oxygenation
no-MODS: patients did not develop MODS
PICU: pediatric intensive care unit
DE: Differentially expressed
FDR: Area under curev
PCA: principal component analysis
OR: Odds ratio

## Supplementary information

**Figure S1.**
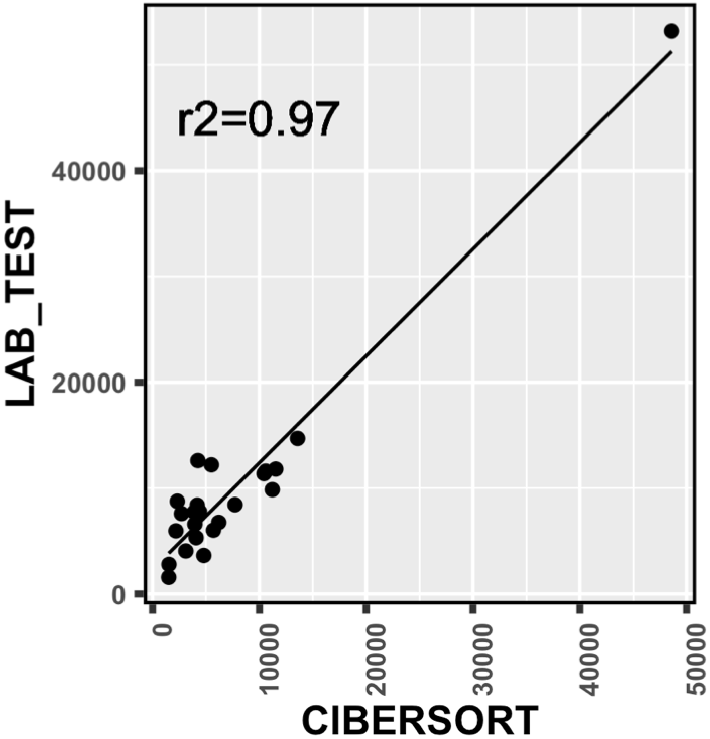
Correlation of neutrophils obtained from lab test data and derived from CIBERSORT. The values closely correlated with each other (Correlation value= 0.97).

**Figure S2.**
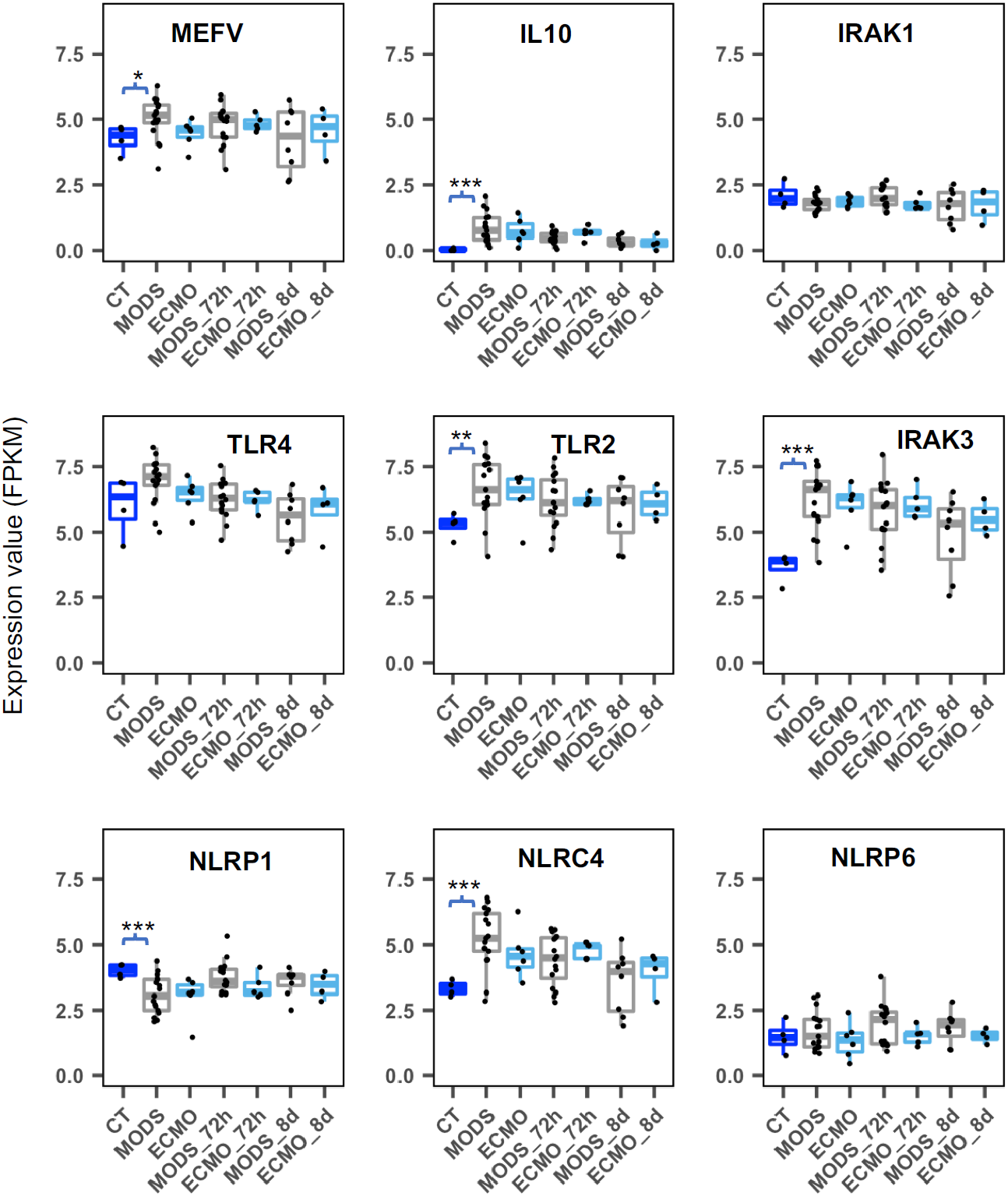
Expression of monocyte genes (based on study of Hall et al., 2007) in control (CT), MODS and ECMO patients at different time points (0h, 72h and 8d). (* 0.01 < P value < 0.05; ** 0.001 < P value < 0.01; *** 7.3e-6 < P value < 0.001).

**Figure S3.**
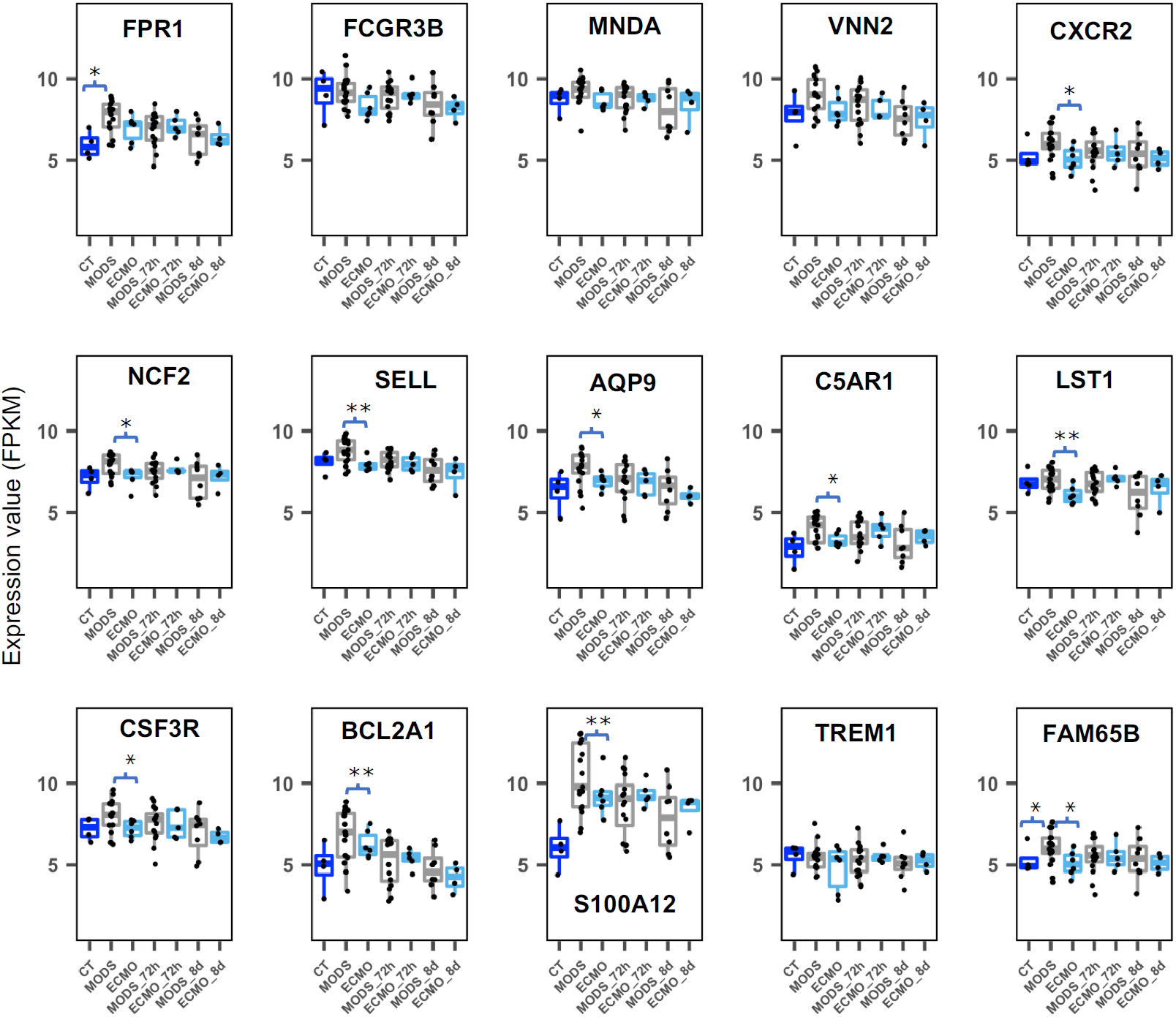
Expression of neutrophils genes (based on CIBERSORT cell marker) in control (CT), MODS and ECMO patients at different time points(0h, 72h and 8d). (* 0.05 > p <= 0.01 and ** 0.01 > p < 0.001).

**Figure S4.**
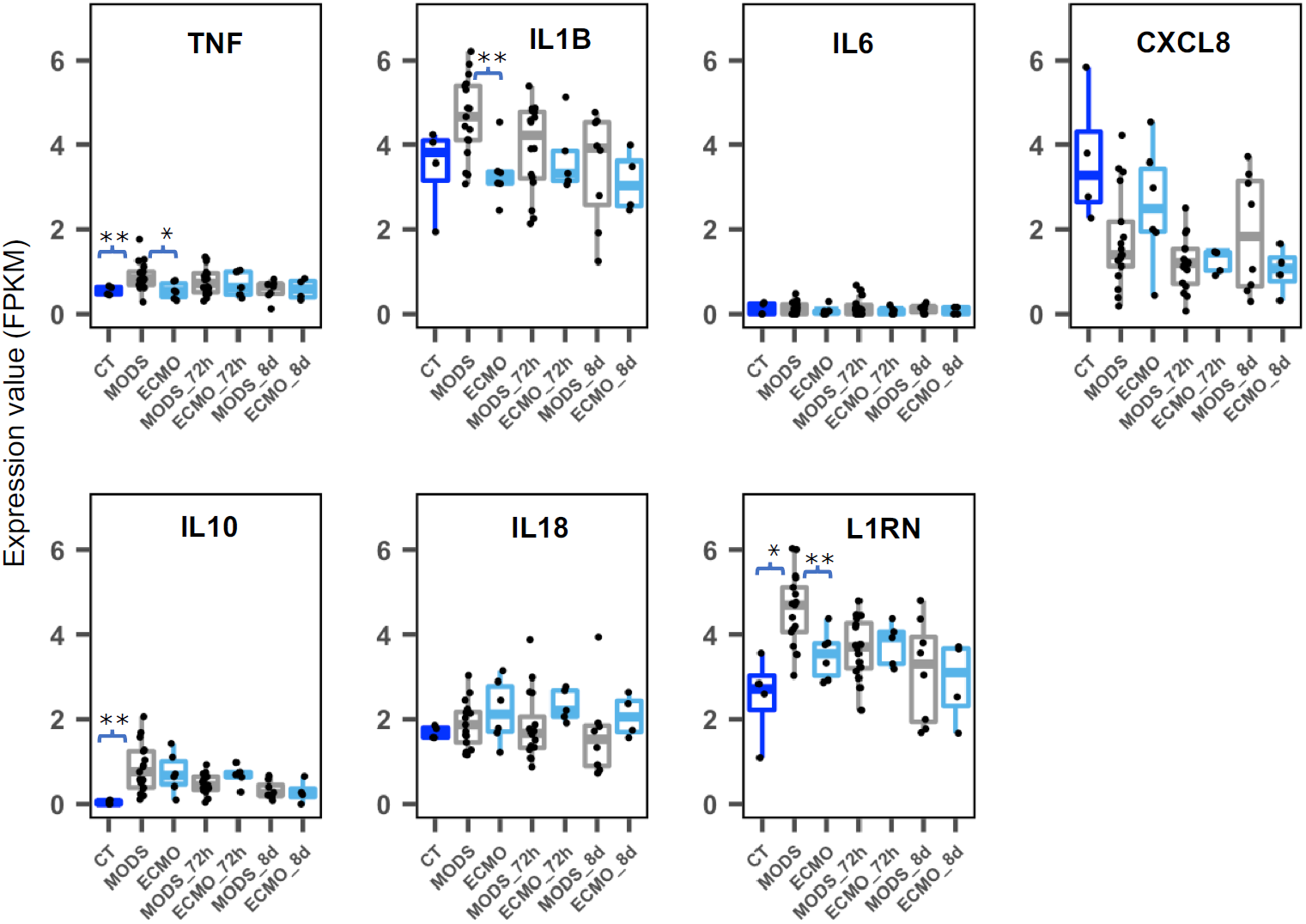
Expression of cytokines genes (based on study of Hall et al., 2007) in control (CT), MODS and ECMO patients at different time points(0h, 72h and 8d). (* 0.05 > p <= 0.01 and ** 0.01 > p < 2.5e-05).

**Figure S5.**
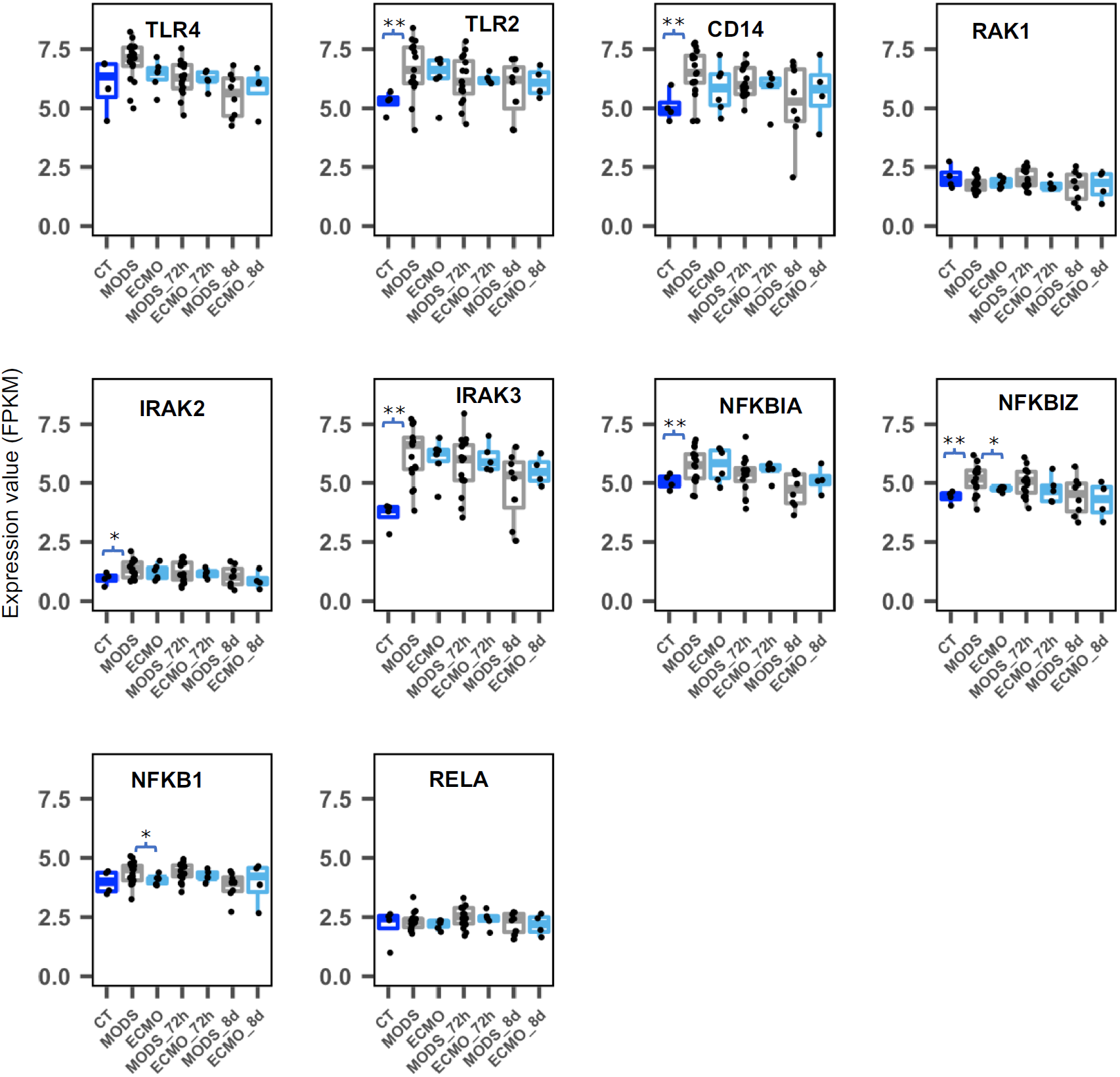
Expression of NF-kB signaling pathway (based on study of Hall et al., 2007) in control (CT), MODS and ECMO patients at different time points(0h, 72h and 8d). (* 0.05 > p <= 0.01 and ** 0.01 > p < 7.3e-05).

**Figure S6.**
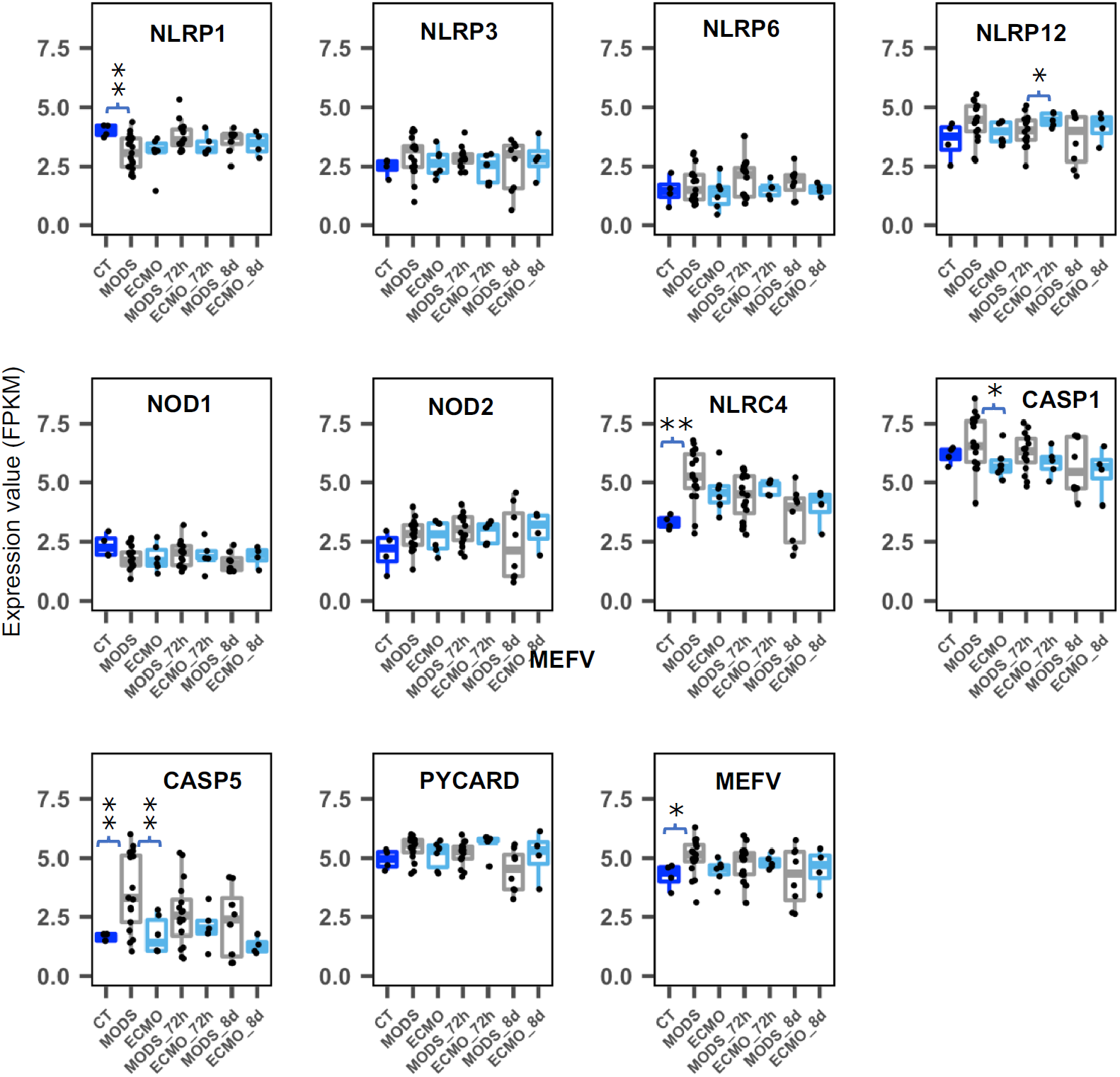
Expression of genes involved in inflammasome elements (based on study of Hall et al., 2007) in control (CT), MODS and ECMO patients at different time points(0h, 72h and 8d). (* 0.05 > p <= 0.01 and ** 0.01 > p < 7.8e-06).

**Figure S7.**
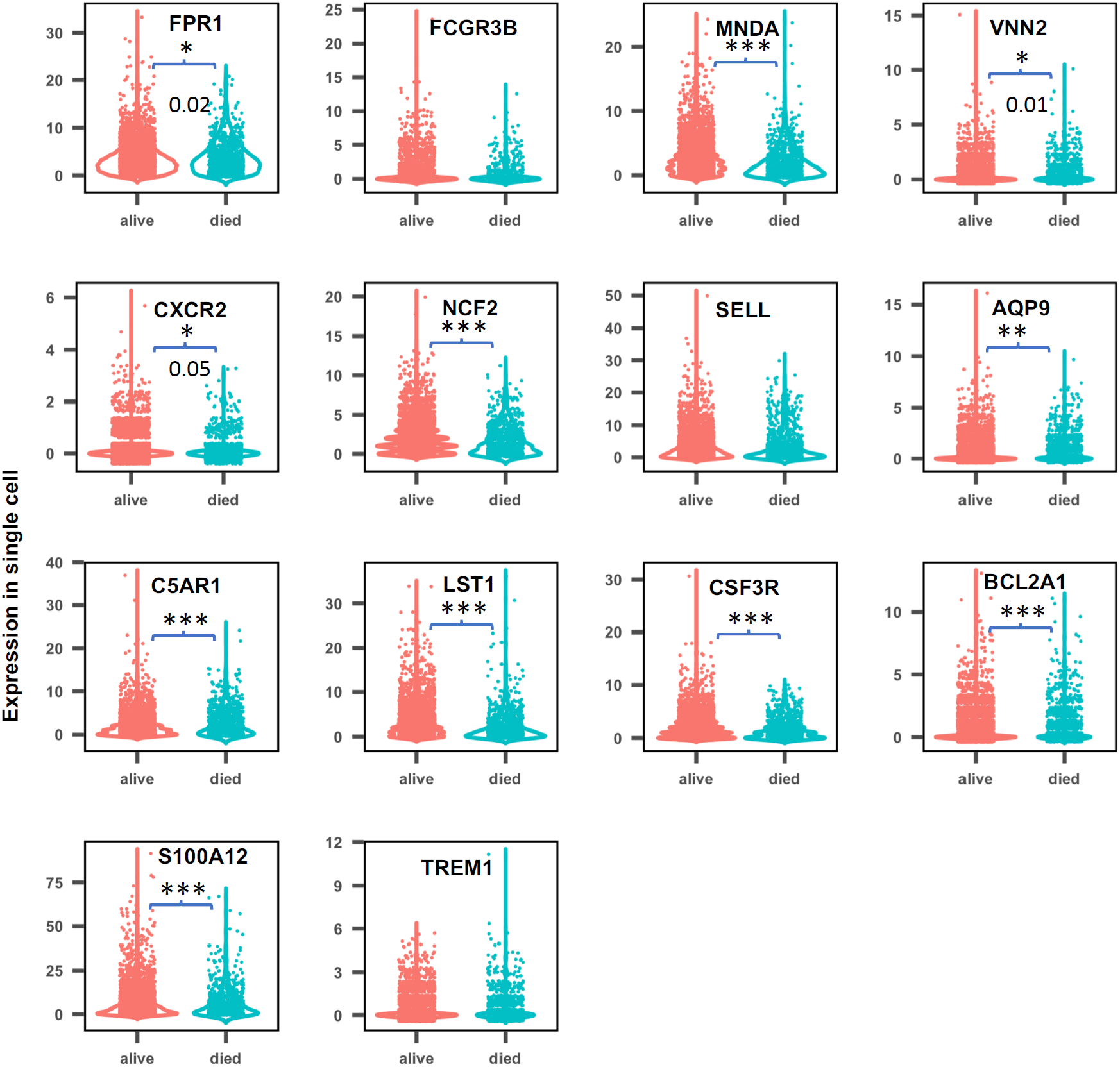
Expression of marker genes for neutrophils cells in single cell data of ECMO adult patients (Kort et al., 2019). Red-Surviving ECMO patients and Green-Died ECMO patients. (* 0.01 < P value < 0.05; ** 0.001 < P value < 0.01; *** 2e-16 < P value < 0.001).

**Figure S8.**
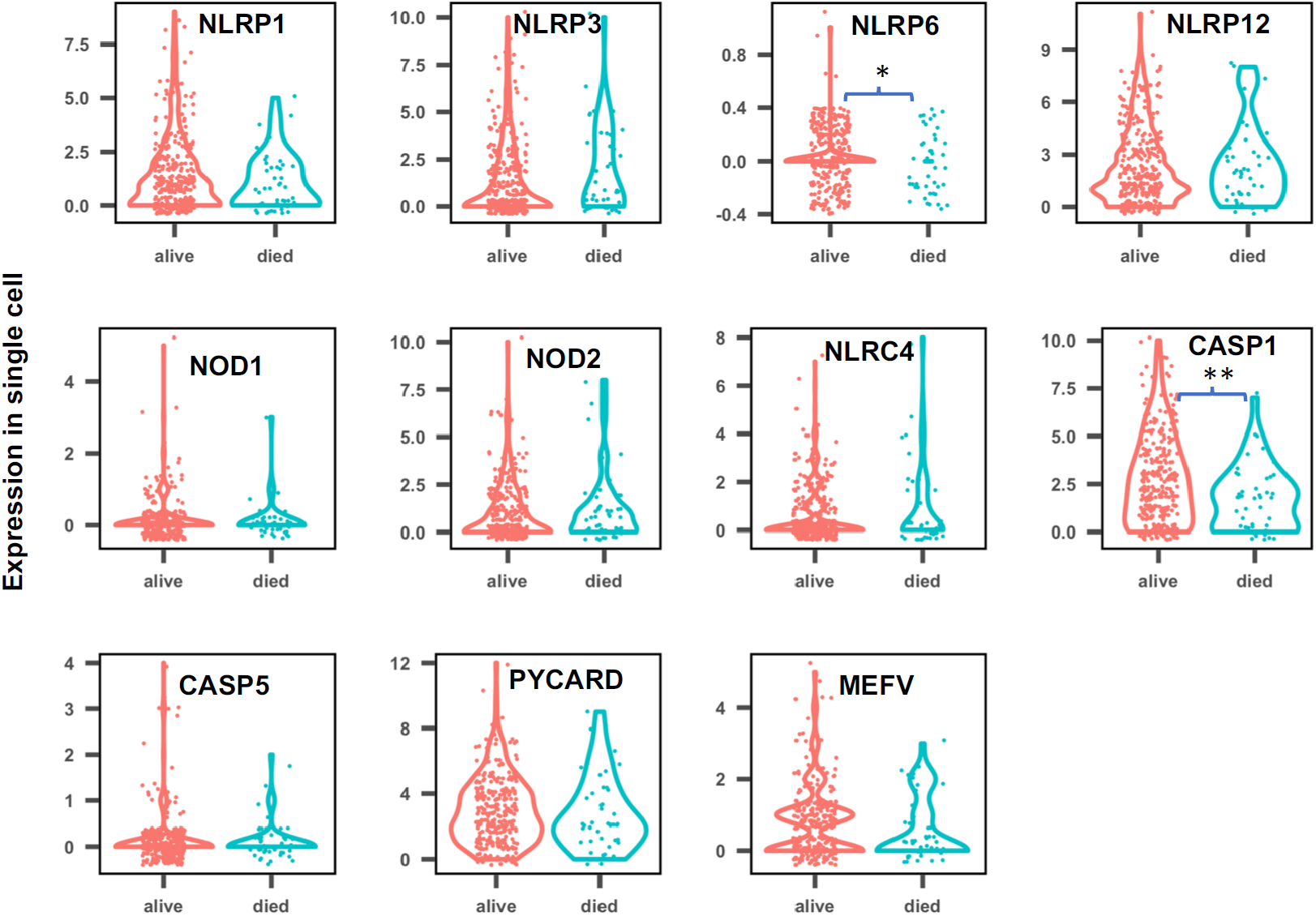
Expression of genes involved in inflammatory response in each cell from the single cell data of ECMO adult patients (Kort et al., 2019). Red-Surviving ECMO patients and Green-Died ECMO patients. (* 0.01 < P value < 0.05 and ** 0.0008 < P value < 0.01.

**Figure S9.**
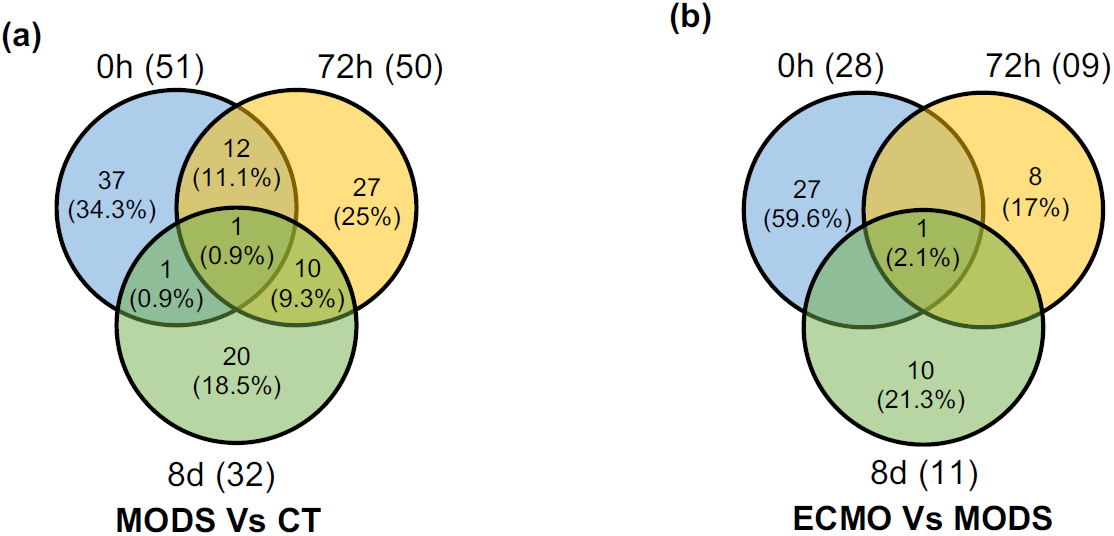
Comparisons of differential gene expression. Venn diagram showing the comparisons of differentially expressed genes in between (a) MODS and control (CT) and (b) in between ECMO and MODS patients at different time points; baseline (0h), 72h and 8d.

**Figure S10.**
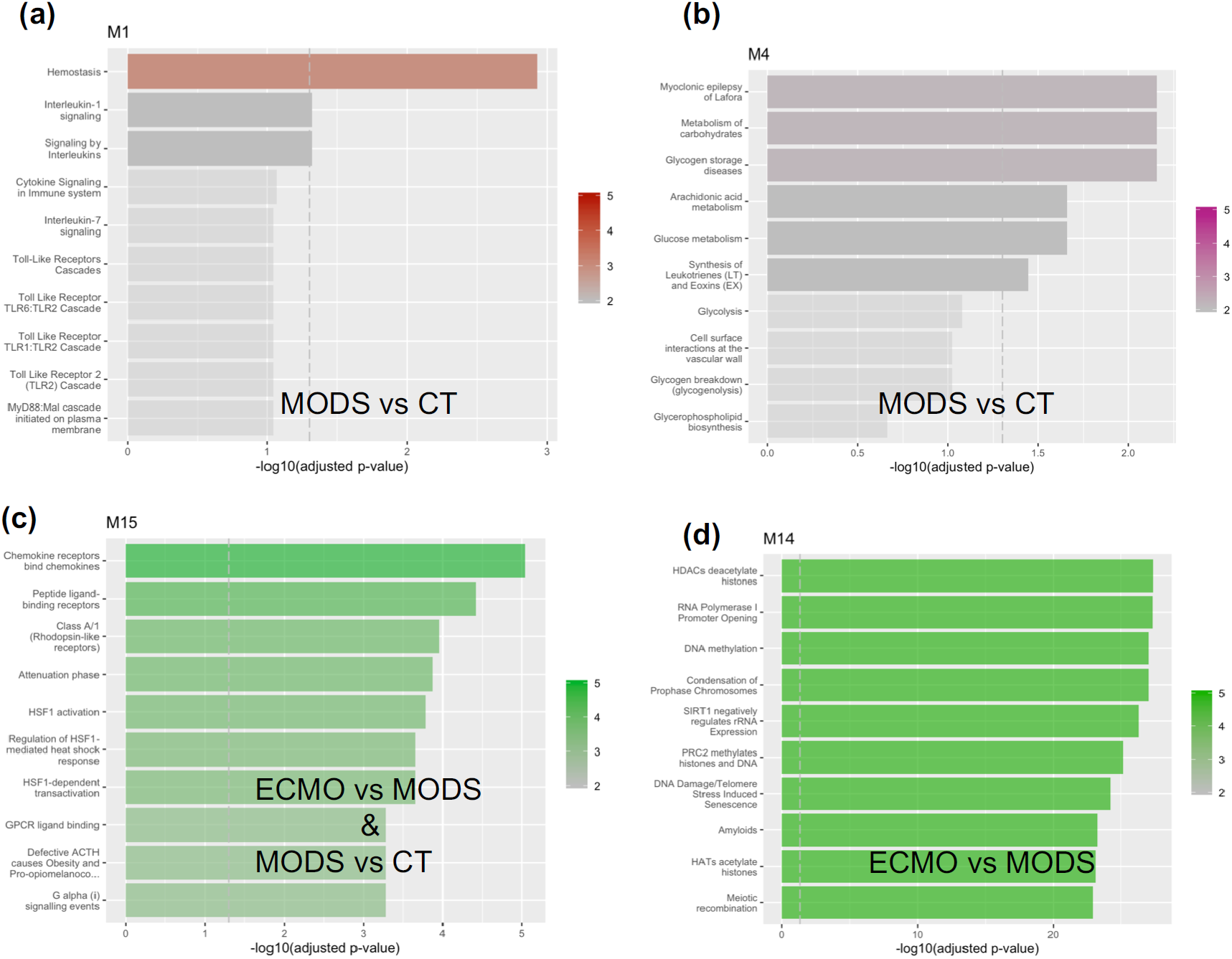
Pathways enriched by the genes present in different co-expressed networks module. (a, b) Enriched pathways are shown by the DE genes from MODS vs CT and mapped on two co-expressed networks. (c) Enriched pathways shared by the DE genes in MODS vs CT and in ECMO vs MODS are shown. This showed the transition from MODS to ECMO. (d) In addition, epigenetic modifications related processes were activated in the severe MODS patients, who require ECMO support.

**Figure S11.**
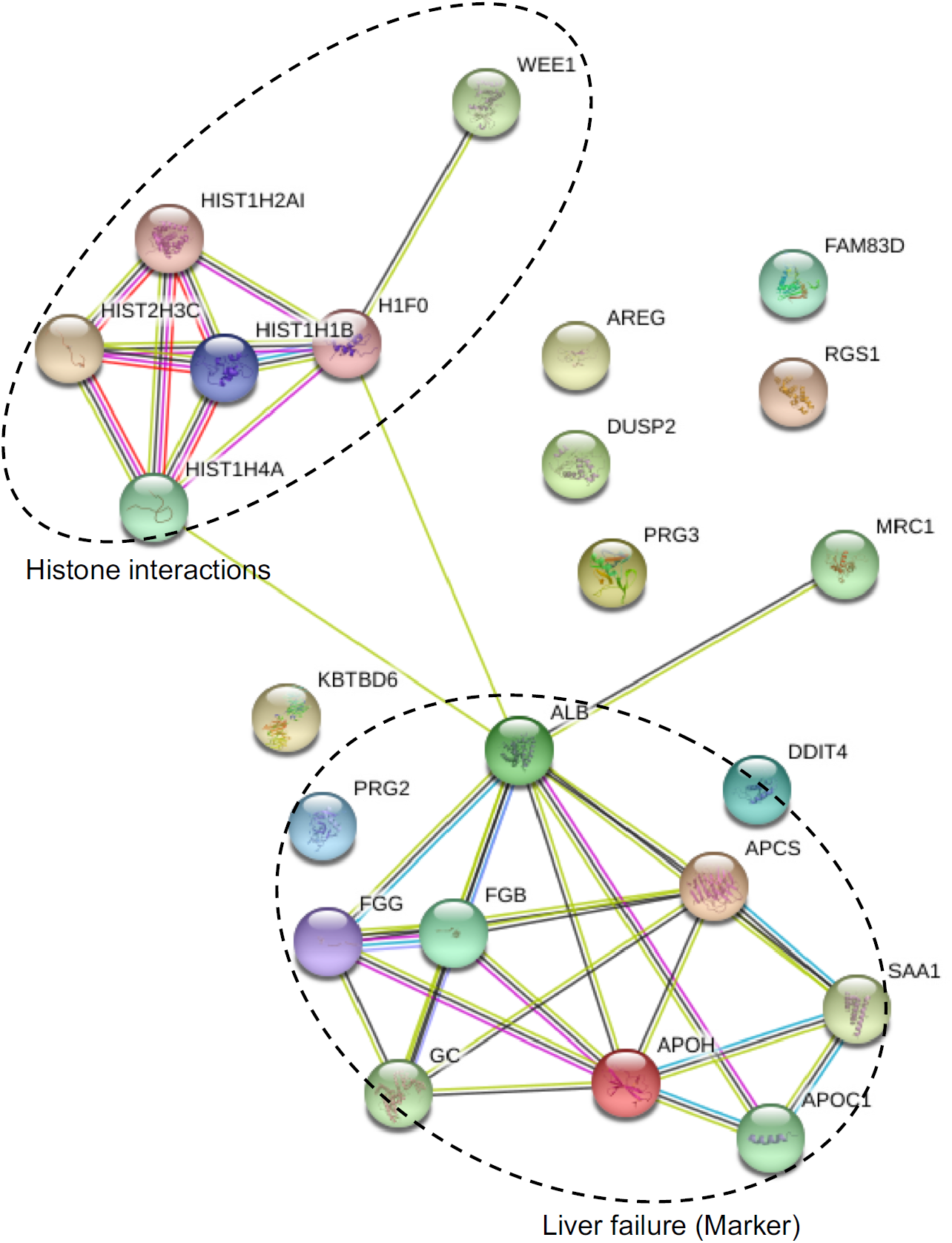
Interaction of genes associated with ECMO at baseline (0h). Two main networks (Histone interactions and gene markers for liver failure) were enriched.

**Figure S12.**
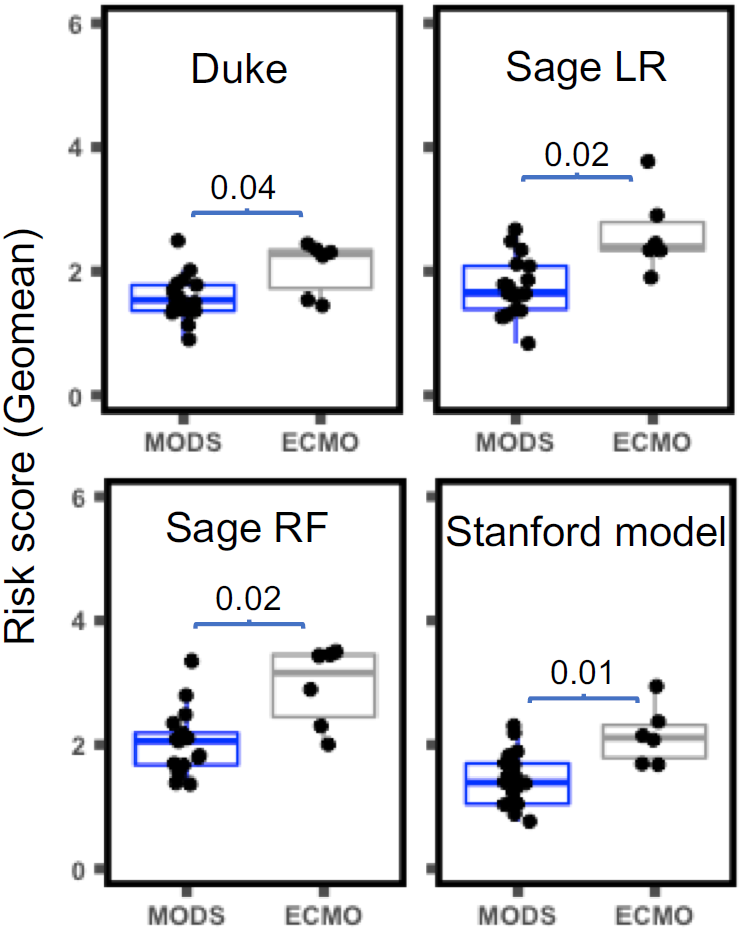
Risk scores derived from the putative signatures predicted for sepsis patients (Sweeney et al., 2018). The signatures have been derived from different models namely, Duke, Sage LR, Sage RF and Stanford. These signatures are composed of two categories, i.e, positively and negatively associated with patients mortality. However, only the signature which are positively associated with patients mortality showed the difference in MODS and ECMO.

**Figure S13.**
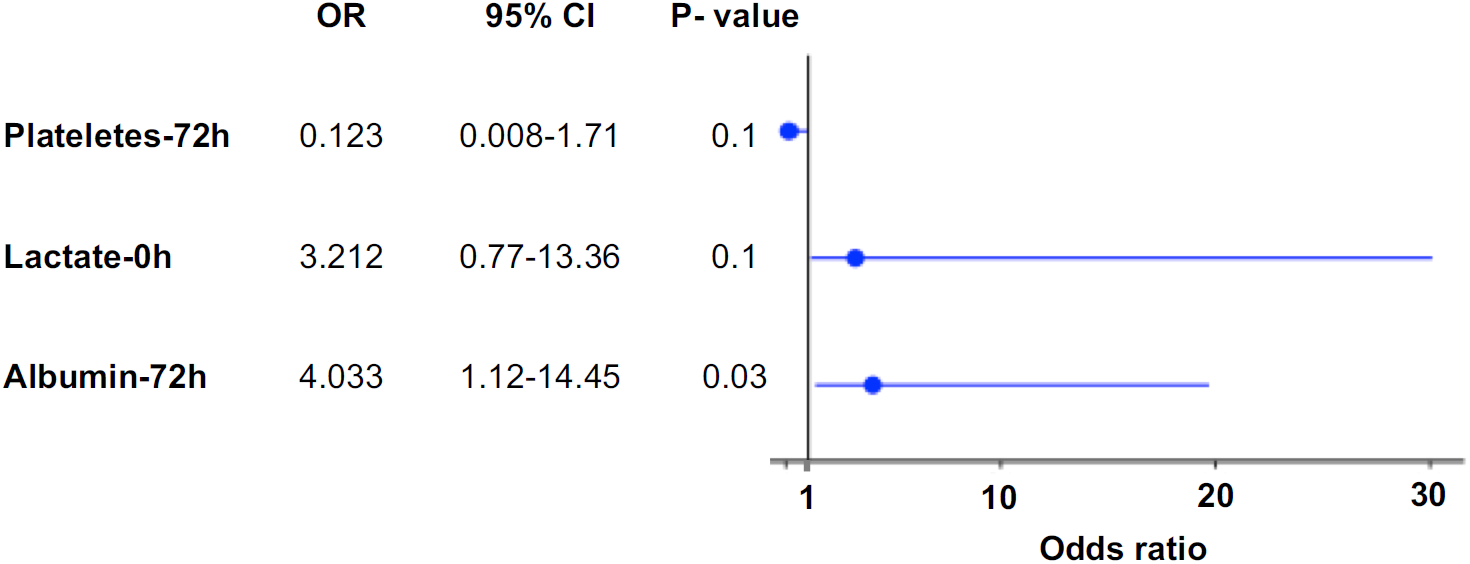
Odds ratio for clinical data. Clinical data available for all the ECMO and MODS patients at different time points were used to compute the odds ratio. Significantly (P value ≤ 0.03) higher odds ratio for Albumin was observed in ECMO as compared to MODS patients.

**Figure S14.**
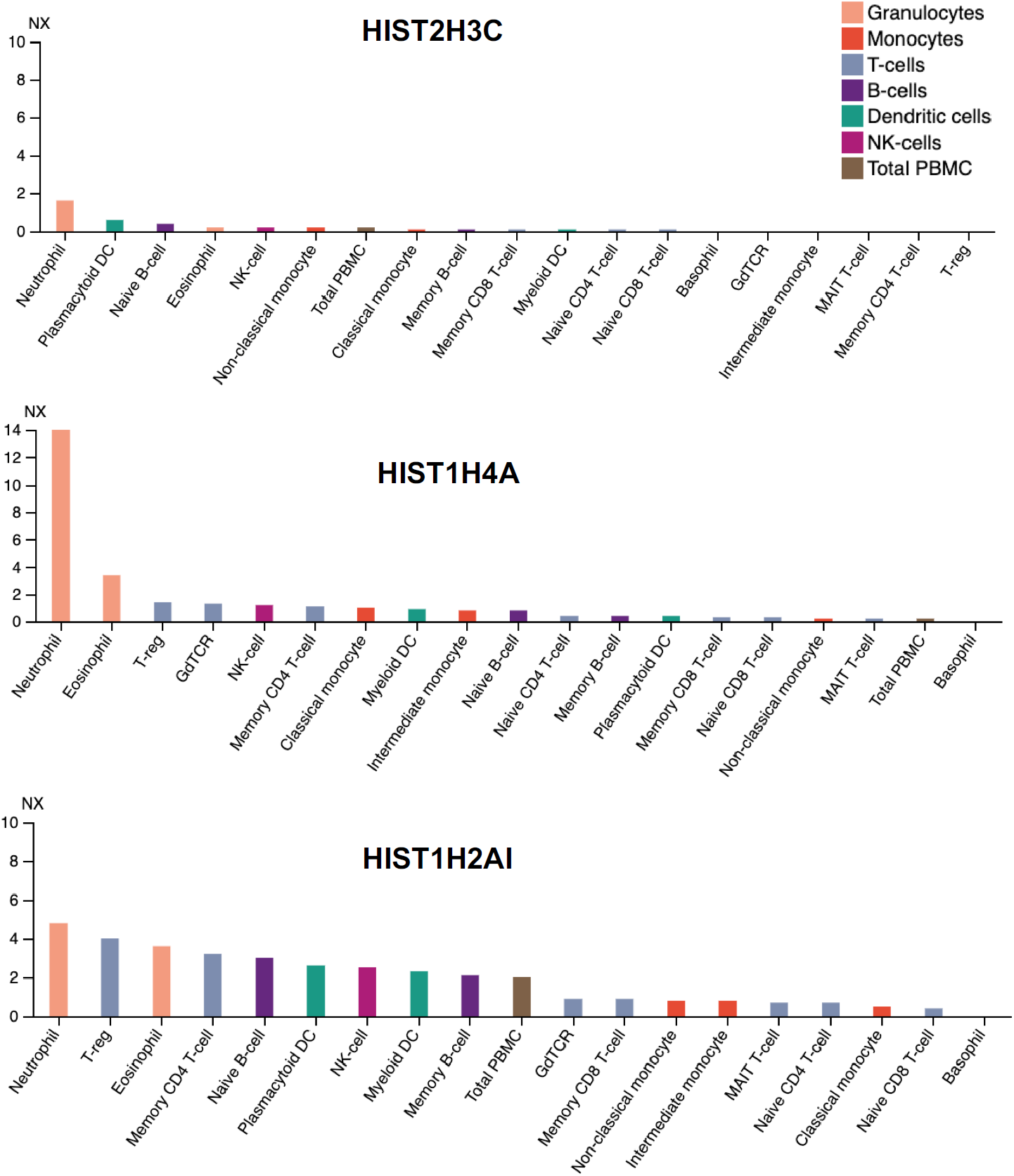
Expression pattern of some of histone genes in blood cells (Human protein atlas). It was observed that HIST2H3C, HIST1H4A, HIST1H2AI is highly expressed in neutrophils.

**Figure S15.**
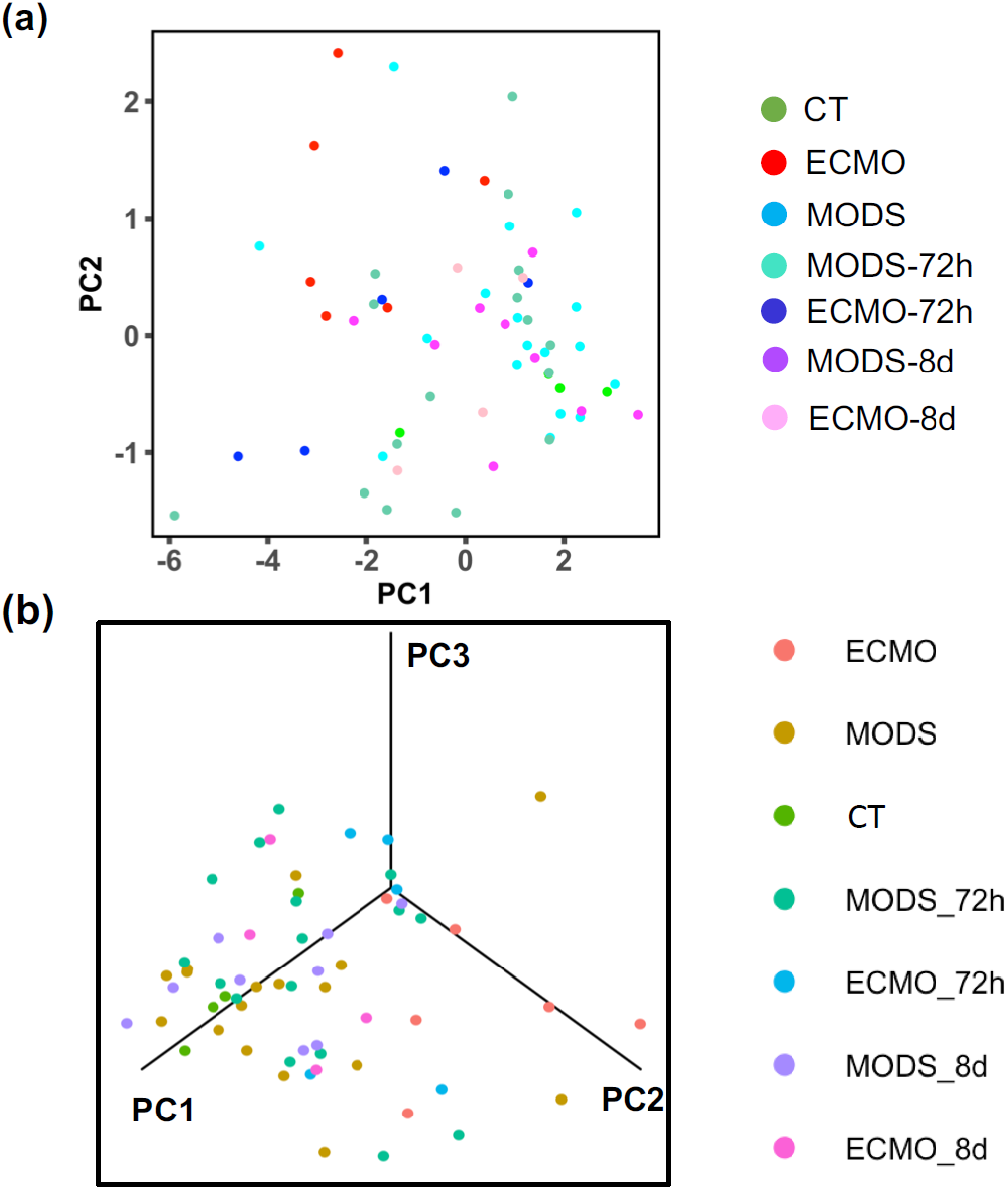
Labeled PCA comparing CT, MODS and ECMO at different time points. (a) Labeled PCA based on gene signature separated all the patients of different time points into CT, MODS and ECMO group. (b) PC1 and PC2 with PC3 provide more clear differentiation of patients.

**Figure S16.**
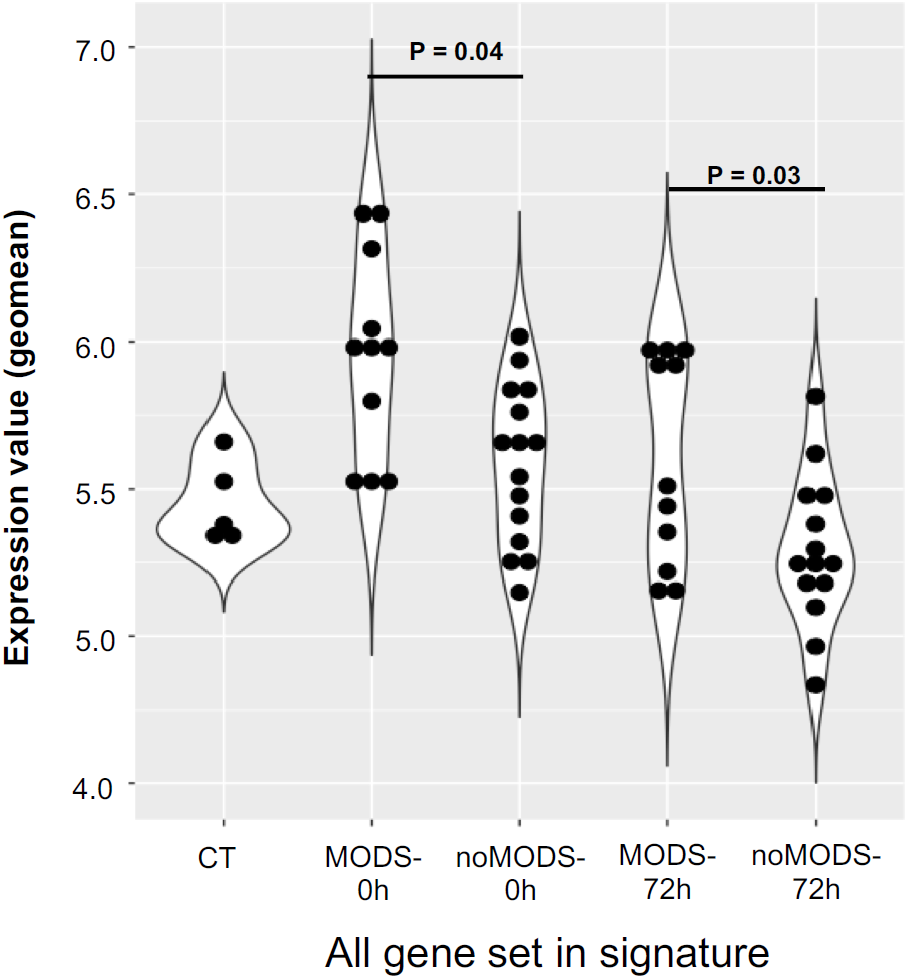
Comparison of all genes in validation data (Cabrera et at., 2017). The violin plot displayed differences in the expression values in MODS and noMODS patients at (P = 0.04) 0h and (P = 0.03) 72h time point.

**Figure S17.**
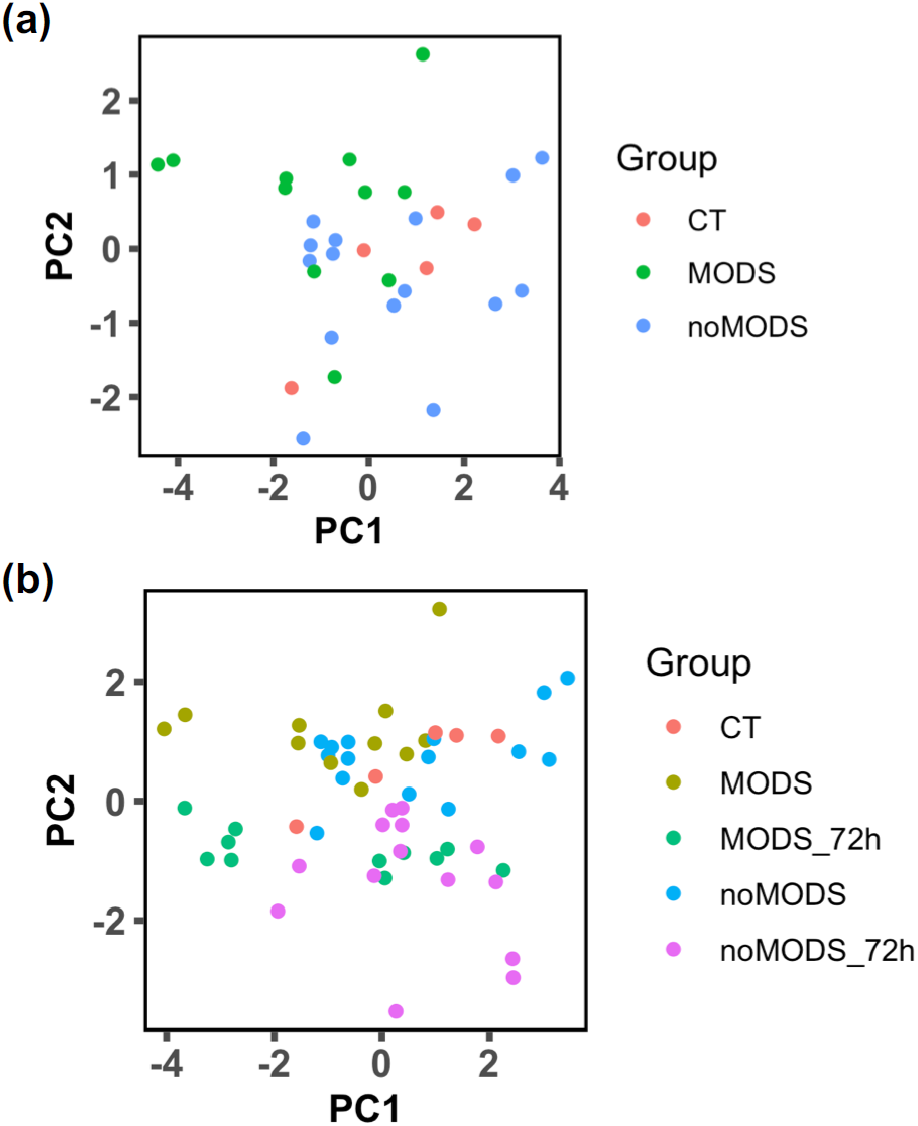
Labeled PCA separated the control (CT), noMODS (patients doesn’t develop MODS) and MODS (develop MODS) patients in the validation cohort (Cabrera et at., 2017). (a) The labeled PCA using the signature genes associated with ECMO separated the MODS and noMODS. (b)The labeled PCA from different time points also separated MODS and noMODS patients with minimal overlap of patients at 72h. Although patients in the validation cohort data were adult, the remarkable separation indirectly validated the signature genes associated with pediatrics ECMO.

**Table S1.**
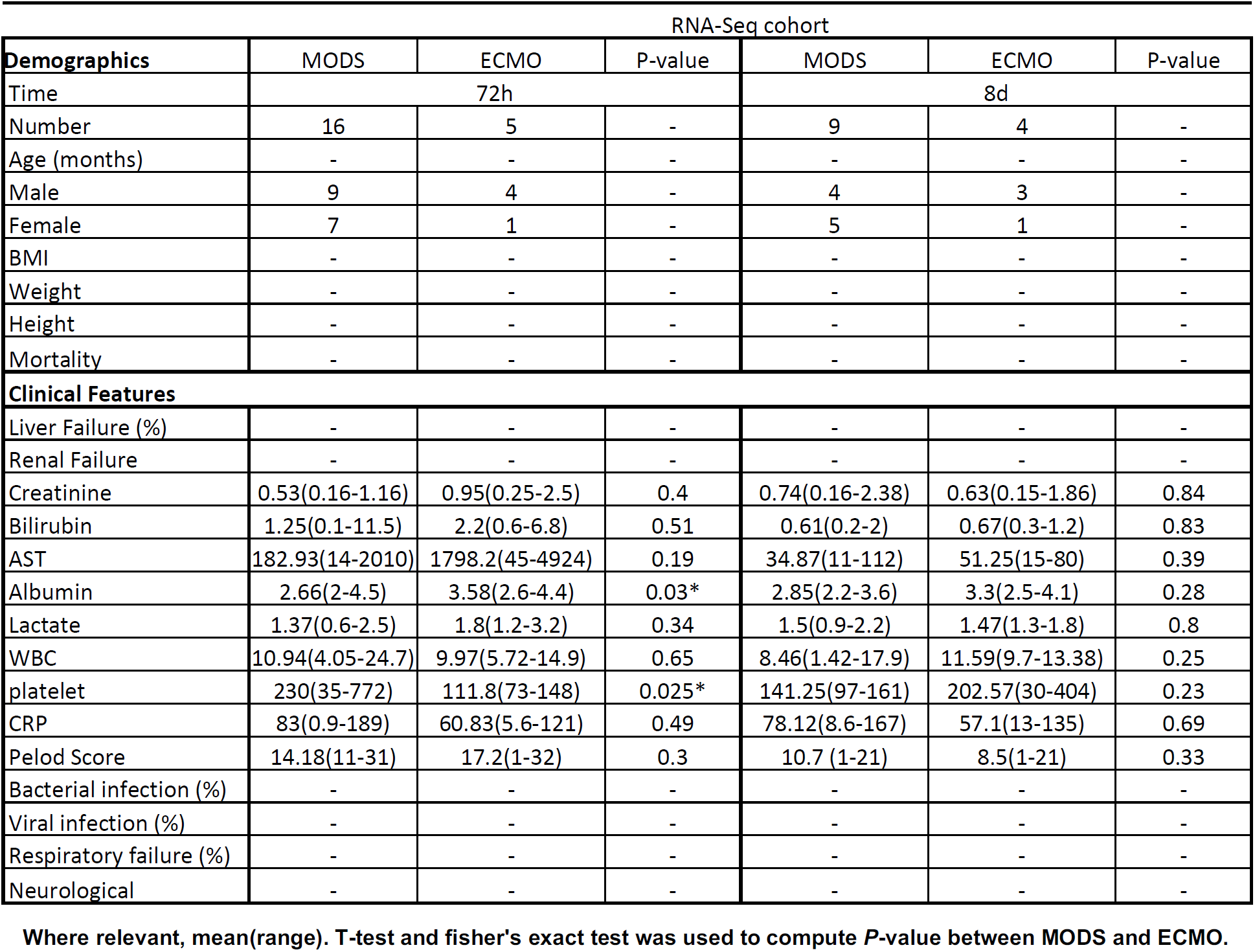
Patient demographics at 72h and 8d time points.

| RNA-Seq cohort |  |  |  |  |  |  |
| --- | --- | --- | --- | --- | --- | --- |
| Demographics | MODS | ECMO | P-value | MODS | ECMO | P-value |
| Time | 72h |  |  | 8d |  |  |
| Number | 16 | 5 | - | 9 | 4 | - |
| Age (months) | - | - | - | - | - | - |
| Male | 9 | 4 | - | 4 | 3 | - |
| Female | 7 | 1 | - | 5 | 1 | - |
| BMI | - | - | - | - | - | - |
| Weight | - | - | - | - | - | - |
| Height | - | - | - | - | - | - |
| Mortality | - | - | - | - | - | - |
| **Clinical Features** |  |  |  |  |  |  |
| Liver Failure (%) | - | - | - | - | - | - |
| Renal Failure | - | - | - | - | - | - |
| Creatinine | 0.53(0.16-1.16) | 0.95(0.25-2.5) | 0.4 | 0.74(0.16-2.38) | 0.63(0.15-1.86) | 0.84 |
| Bilirubin | 1.25(0.1-11.5) | 2.2(0.6-6.8) | 0.51 | 0.61(0.2-2) | 0.67(0.3-1.2) | 0.83 |
| AST | 182.93(14-2010) | 1798.2(45-4924) | 0.19 | 34.87(11-112) | 51.25(15-80) | 0.39 |
| Albumin | 2.66(2-4.5) | 3.58(2.6-4.4) | 0.03* | 2.85(2.2-3.6) | 3.3(2.5-4.1) | 0.28 |
| Lactate | 1.37(0.6-2.5) | 1.8(1.2-3.2) | 0.34 | 1.5(0.9-2.2) | 1.47(1.3-1.8) | 0.8 |
| WBC | 10.94(4.05-24.7) | 9.97(5.72-14.9) | 0.65 | 8.46(1.42-17.9) | 11.59(9.7-13.38) | 0.25 |
| platelet | 230(35-772) | 111.8(73-148) | 0.025* | 141.25(97-161) | 202.57(30-404) | 0.23 |
| CRP | 83(0.9-189) | 60.83(5.6-121) | 0.49 | 78.12(8.6-167) | 57.1(13-135) | 0.69 |
| Pelod Score | 14.18(11-31) | 17.2(1-32) | 0.3 | 10.7 (1-21) | 8.5(1-21) | 0.33 |
| Bacterial infection (%) | - | - | - | - | - | - |
| Viral infection (%) | - | - | - | - | - | - |
| Respiratory failure (%) | - | - | - | - | - | - |
| Neurological | - | - | - | - | - | - |

Where relevant, mean(range). T-test and fisher's exact test was used to compute *P*-value between MODS and ECMO.

**Table S2.** List of differentially expressed genes in ECMO as compared to MODS patients at baseline (0h).

| Gene | Ensemble | Log<sup>2</sup> Fold Change | Log CPM | P-value | Adj. P-value | Protein coding | Function |
| --- | --- | --- | --- | --- | --- | --- | --- |
| RGS1 | ENSG00000090104 | 4.12 | 2.75 | 1.3E-08 | 0.001 | Y | regulator of G-protein signaling 1 |
| APOH | ENSG00000091583 | 6.91 | -2.00 | 2.5E-08 | 0.001 | Y | apolipoprotein H (beta-2-glycoprotein I) |
| FAM83D | ENSG00000101447 | 1.97 | -0.23 | 1.7E-07 | 0.010 | Y | family with sequence similarity 83, member D |
| AREG | ENSG00000109321 | 3.51 | 0.49 | 2.0E-06 | 0.118 | Y | amphiregulin |
| APOC1 | ENSG00000130208 | 3.37 | -1.93 | 2.0E-06 | 0.121 | Y | apolipoprotein C-I |
| APCS | ENSG00000132703 | 5.12 | -2.93 | 5.6E-07 | 0.034 | Y | amyloid P component, serum |
| GC | ENSG00000145321 | 6.76 | -2.08 | 3.0E-08 | 0.002 | Y | group-specific component (vitamin D binding protein) |
| PRG3 | ENSG00000156575 | 5.74 | -1.27 | 5.9E-14 | 0.000 | Y | proteoglycan 3 |
| DUSP2 | ENSG00000158050 | 3.13 | 1.79 | 3.4E-08 | 0.002 | Y | dual specificity phosphatase 2 |
| ALB | ENSG00000163631 | 7.92 | 1.80 | 3.6E-09 | 0.000 | Y | albumin |
| KBTBD6 | ENSG00000165572 | 1.69 | 3.50 | 1.2E-07 | 0.007 | Y | kelch repeat and BTB (POZ) domain containing 6 |
| WEE1 | ENSG00000166483 | 1.54 | 2.52 | 1.5E-06 | 0.091 | Y | WEE1 homolog (S. pombe) |
| DDIT4 | ENSG00000168209 | 2.02 | 3.74 | 4.4E-07 | 0.026 | Y | DNA-damage-inducible transcript 4 |
| FGG | ENSG00000171557 | 6.02 | -1.45 | 2.1E-07 | 0.013 | Y | fibrinogen gamma chain |
| FGB | ENSG00000171564 | 7.08 | -1.11 | 6.3E-08 | 0.004 | Y | fibrinogen beta chain |
| SAA1 | ENSG00000173432 | 7.55 | -1.02 | 5.7E-08 | 0.003 | Y | serum amyloid A1 |
| HIST1H1B | ENSG00000184357 | 1.92 | 4.24 | 2.4E-06 | 0.147 | Y | histone cluster 1, H1b |
| PRG2 | ENSG00000186652 | 4.96 | 1.67 | 2.3E-08 | 0.001 | Y | proteoglycan 2 |
| H1F0 | ENSG00000189060 | 1.73 | 4.50 | 3.5E-06 | 0.212 | Y | H1 histone family, member 0 |
| HIST1H2AI | ENSG00000196747 | 1.62 | 3.86 | 3.4E-06 | 0.207 | Y | histone cluster 1, H2ai |
| RNA5S9 | ENSG00000201321 | 5.54 | -0.35 | 5.7E-08 | 0.003 | Y | RNA, 5S ribosomal 9 |
| HIST2H3C | ENSG00000203811 | 2.11 | 3.88 | 9.1E-07 | 0.055 | Y | histone cluster 2, H3c |
| RNU1-67P | ENSG00000207175 | 3.94 | 0.78 | 1.2E-08 | 0.001 | N | NA |
| HMGB1P30 | ENSG00000244089 | 3.59 | -2.84 | 5.2E-08 | 0.003 | Y | high mobility group box 1 pseudogene 30 |
| RP11-1H15.1 | ENSG00000254765 | 3.83 | -3.28 | 2.1E-06 | 0.125 | N | NA |
| MRC1 | ENSG00000260314 | 3.55 | 2.13 | 3.3E-08 | 0.002 | Y | mannose receptor, C type 1 |
| CTD-2033D15.2 | ENSG00000276107 | 3.32 | -0.61 | 1.1E-08 | 0.001 | N | NA |
| HIST1H4A | ENSG00000278637 | 1.85 | 0.24 | 1.0E-06 | 0.061 | Y | histone cluster 1, H4a |

**Table S3.**
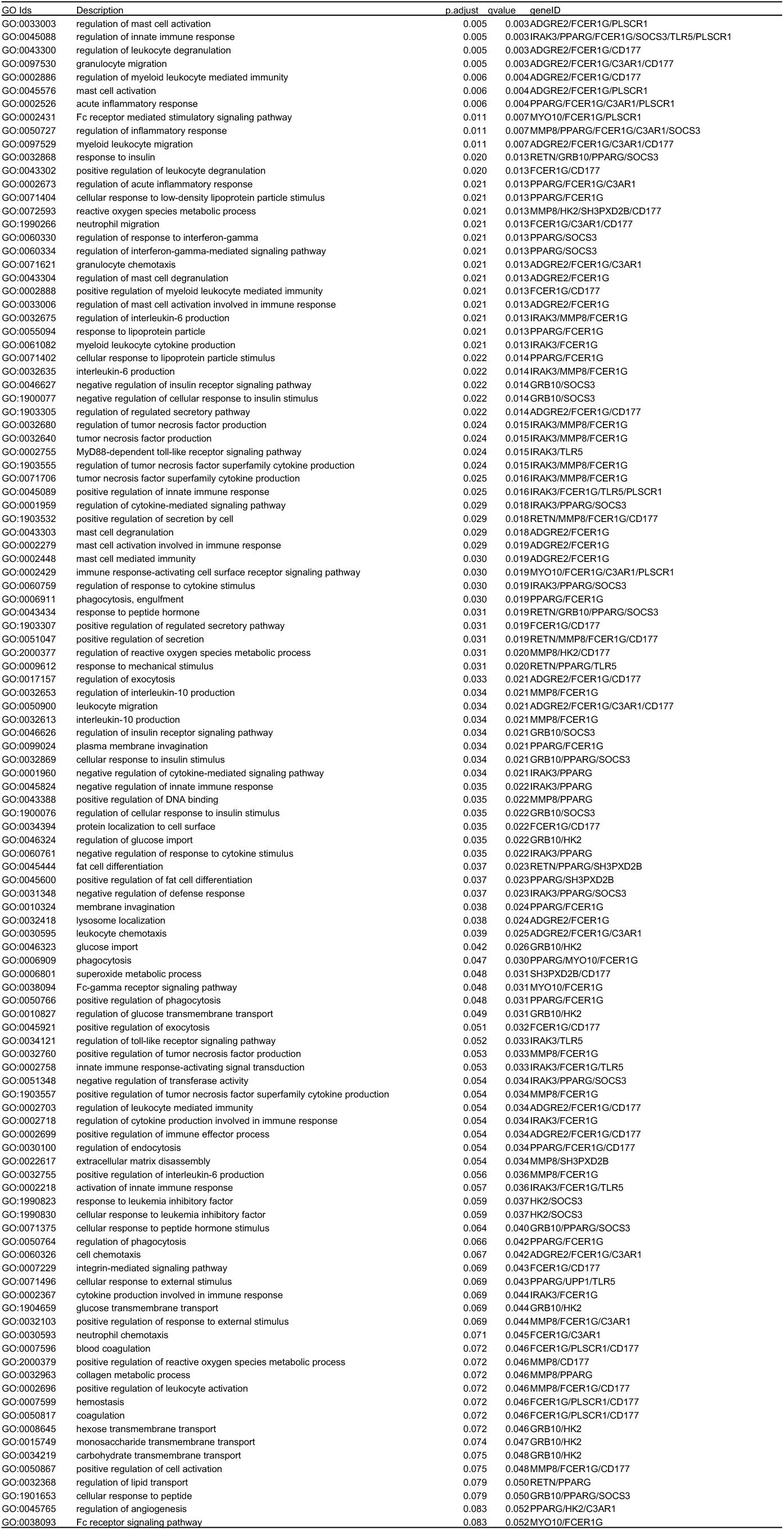
Complete list of gene enriched in different processes in MODS as compared to CT at base line (0h).

**Table S4.** Complete list of gene enriched in different processes in ECMO as compared to MODS at base line (0h).

| GO Id | Description | p.adjust | qvalue | geneID |
| --- | --- | --- | --- | --- |
| GO:0006342 | chromatin silencing | 0.00 | 0.00 | HIST1H1B/H1F0/HIST2H3C/HIST2H3A/HIST1H4A |
| GO:0060968 | regulation of gene silencing | 0.00 | 0.00 | HIST1H1B/H1F0/HIST2H3C/HIST2H3A/HIST1H4A |
| GO:0045814 | negative regulation of gene expression, epigenetic | 0.00 | 0.00 | HIST1H1B/H1F0/HIST2H3C/HIST2H3A/HIST1H4A |
| GO:0006334 | nucleosome assembly | 0.00 | 0.00 | HIST1H1B/H1F0/HIST2H3C/HIST2H3A/HIST1H4A |
| GO:0031497 | chromatin assembly | 0.00 | 0.00 | HIST1H1B/H1F0/HIST2H3C/HIST2H3A/HIST1H4A |
| GO:0061045 | negative regulation of wound healing | 0.00 | 0.00 | APOH/APCS/FGG/FGB |
| GO:0007596 | blood coagulation | 0.00 | 0.00 | APOH/FGG/FGB/SAA1/HIST2H3C/HIST2H3A |
| GO:0031639 | plasminogen activation | 0.00 | 0.00 | APOH/FGG/FGB |
| GO:0034728 | nucleosome organization | 0.00 | 0.00 | HIST1H1B/H1F0/HIST2H3C/HIST2H3A/HIST1H4A |
| GO:0007599 | hemostasis | 0.00 | 0.00 | APOH/FGG/FGB/SAA1/HIST2H3C/HIST2H3A |
| GO:0050817 | coagulation | 0.00 | 0.00 | APOH/FGG/FGB/SAA1/HIST2H3C/HIST2H3A |
| GO:1903035 | negative regulation of response to wounding | 0.00 | 0.00 | APOH/APCS/FGG/FGB |
| GO:0006333 | chromatin assembly or disassembly | 0.00 | 0.00 | HIST1H1B/H1F0/HIST2H3C/HIST2H3A/HIST1H4A |
| GO:0072378 | blood coagulation, fibrin clot formation | 0.00 | 0.00 | APOH/FGG/FGB |
| GO:0042730 | fibrinolysis | 0.00 | 0.00 | APOH/FGG/FGB |
| GO:0006323 | DNA packaging | 0.00 | 0.00 | HIST1H1B/H1F0/HIST2H3C/HIST2H3A/HIST1H4A |
| GO:0072376 | protein activation cascade | 0.00 | 0.00 | APOH/APCS/FGG/FGB |
| GO:0000183 | chromatin silencing at rDNA | 0.00 | 0.00 | HIST2H3C/HIST2H3A/HIST1H4A |
| GO:0065004 | protein-DNA complex assembly | 0.00 | 0.00 | HIST1H1B/H1F0/HIST2H3C/HIST2H3A/HIST1H4A |
| GO:0002576 | platelet degranulation | 0.00 | 0.00 | APOH/ALB/FGG/FGB |
| GO:0061041 | regulation of wound healing | 0.00 | 0.00 | APOH/APCS/FGG/FGB |
| GO:0071103 | DNA conformation change | 0.00 | 0.00 | HIST1H1B/H1F0/HIST2H3C/HIST2H3A/HIST1H4A |
| GO:0030195 | negative regulation of blood coagulation | 0.00 | 0.00 | APOH/FGG/FGB |
| GO:1900047 | negative regulation of hemostasis | 0.00 | 0.00 | APOH/FGG/FGB |
| GO:0071824 | protein-DNA complex subunit organization | 0.00 | 0.00 | HIST1H1B/H1F0/HIST2H3C/HIST2H3A/HIST1H4A |
| GO:0031638 | zymogen activation | 0.00 | 0.00 | APOH/FGG/FGB |
| GO:0050819 | negative regulation of coagulation | 0.00 | 0.00 | APOH/FGG/FGB |
| GO:1903034 | regulation of response to wounding | 0.00 | 0.00 | APOH/APCS/FGG/FGB |
| GO:0032102 | negative regulation of response to external stimulus | 0.00 | 0.00 | APOH/APCS/FGG/FGB/SAA1 |
| GO:0030193 | regulation of blood coagulation | 0.00 | 0.00 | APOH/FGG/FGB |
| GO:0045652 | regulation of megakaryocyte differentiation | 0.00 | 0.00 | HIST2H3C/HIST2H3A/HIST1H4A |
| GO:1900046 | regulation of hemostasis | 0.00 | 0.00 | APOH/FGG/FGB |
| GO:0050818 | regulation of coagulation | 0.00 | 0.00 | APOH/FGG/FGB |
| GO:0016584 | nucleosome positioning | 0.00 | 0.00 | HIST1H1B/H1F0 |
| GO:0060964 | regulation of gene silencing by miRNA | 0.00 | 0.00 | HIST2H3C/HIST2H3A/HIST1H4A |
| GO:0060147 | regulation of posttranscriptional gene silencing | 0.00 | 0.00 | HIST2H3C/HIST2H3A/HIST1H4A |
| GO:0060966 | regulation of gene silencing by RNA | 0.00 | 0.00 | HIST2H3C/HIST2H3A/HIST1H4A |
| GO:0030219 | megakaryocyte differentiation | 0.00 | 0.00 | HIST2H3C/HIST2H3A/HIST1H4A |
| GO:0031936 | negative regulation of chromatin silencing | 0.00 | 0.00 | HIST1H1B/H1F0 |
| GO:0051004 | regulation of lipoprotein lipase activity | 0.01 | 0.00 | APOH/APOC1 |
| GO:0045637 | regulation of myeloid cell differentiation | 0.01 | 0.00 | APCS/HIST2H3C/HIST2H3A/HIST1H4A |
| GO:0034375 | high-density lipoprotein particle remodeling | 0.01 | 0.00 | APOC1/ALB |
| GO:0006898 | receptor-mediated endocytosis | 0.01 | 0.01 | APOC1/ALB/SAA1/MRC1 |
| GO:0031935 | regulation of chromatin silencing | 0.01 | 0.01 | HIST1H1B/H1F0 |
| GO:0034116 | positive regulation of heterotypic cell-cell adhesion | 0.01 | 0.01 | FGG/FGB |
| GO:0038111 | interleukin-7-mediated signaling pathway | 0.01 | 0.01 | HIST2H3C/HIST2H3A |
| GO:0045907 | positive regulation of vasoconstriction | 0.01 | 0.01 | FGG/FGB |
| GO:0098760 | response to interleukin-7 | 0.01 | 0.01 | HIST2H3C/HIST2H3A |
| GO:0098761 | cellular response to interleukin-7 | 0.01 | 0.01 | HIST2H3C/HIST2H3A |

## References

1. Weiss SL, Fitzgerald JC, Pappachan J, Wheeler D, Jaramillo-Bustamante JC, Salloo A, et al. Global epidemiology of pediatric severe sepsis: the sepsis prevalence, outcomes, and therapies study. Am J Respir Crit Care Med. 2015; 191(10): 1147–57. https://doi.org/10.1164/rccm.201412-2323OC PMID: 25734408.

2. Bestati N, Leteurtre S, Duhamel A, Proulx F, Grandbastien B, Lacroix J, et al. Differences in organ dysfunctions between neonates and older children: a prospective, observational, multicenter study. Crit Care. 2010; 14(6): R202. https://doi.org/10.1186/cc9323 PMID: 21062434.

3. Jenks CL, Raman L and Dalton HJ. Pediatric extracorporeal membrane oxygenation. Crit Care Clin. 2017; 33(4):825–41. https://doi.org/10.1016/j.ccc.2017.06.005 PMID: 28887930.

4. Sweeney TE, Perumal TM, Henao R, Nichols M, Howrylak JA, Choi AM, et al. A community approach to mortality prediction in sepsis via gene expression analysis. Nat Commun. 2018; 9(1): 694. https://doi.org/10.1038/s41467-018-03078-2 PMID: 29449546.

5. Sweeney TE, Azad TD, Donato M, Haynes WA, Perumal TM, Henao R et al. Unsupervised analysis of transcriptomics in bacterial sepsis across multiple datasets reveals three tobust clusters. Crit Care Med. 2018; 46(6): 915–925. https://doi.org/10.1097/CCM.0000000000003084 PMID: 29537985.

6. Sweeney TE, Shidham A, Wong HR and Khatri P. A comprehensive time-course-based multicohort analysis of sepsis and sterile inflammation reveals a robust diagnostic gene set. Sci Transl Med. 2015; 7(287): 287ra71. https://doi.org/10.1126/scitranslmed.aaa5993 PMID: 25972003.

7. Cabrera CP, Manson J, Shepherd JM, Torrance HD, Watson D, Longhi MP, et al. Signatures of inflammation and impending multiple organ dysfunction in the hyperacute phase of trauma: A prospective cohort study. PLoS Med. 2017; 14(7): e1002352. https://doi.org/10.1371/journal PMID.1002352.

8. Kort EJ, Weiland M, Grins E, Eugster E, Milliron HY, Kelty C, et al. Single cell transcriptomics is a robust approach to defining disease biology in complex clinical settings. bioRxiv. 2019; 568659. https://doi.org/10.1101/568659.

9. Dobin A, Davis CA, Schlesinger F, Drenkow J, Zaleski C, Jha S, et al. STAR: ultrafast universal RNA-seq aligner. Bioinformatics. 2013; 29(1): 15–21. https://doi.org/10.1093/bioinformatics/bts635 PMID:23104886.

10. Robinson MD, McCarthy DJ, Smyth GK. edgeR: a Bioconductor package for differential expression analysis of digital gene expression data. Bioinformatics. 2010; 26(1): 139–40. https://doi.org/10.1093/bioinformatics/btp616 PMID:19910308.

11. Russo PdST, Ferreira GR, Cardozo LE, Burger MC, Arias-Carrasco R, Maruyama SR et al. “CEMiTool: a Bioconductor package for performing comprehensive modular co-expression analyses.” BMC Bioinformatics. 2018: 19(56): 1–13. https://doi.org/10.1186/s12859-018-2053-1 PMID: 29458351.

12. Yu G, Wang L, Han Y and He Q. clusterProfiler: an R package for comparing biological themes among gene clusters. OMICS. 2012: 16(5): 284–287. https://doi.org/10.1089/omi.2011.0118 PMID:22455463.

13. Newman AM, Liu CL, Green MR, Gentles AJ, Feng W, Xu Y, et al. Robust enumeration of cell subsets from tissue expression profiles. Nat Methods. 2015; 12(5): 453. https://doi.org/10.1038/nmeth.3337 PMID: 25822800.

14. Schölkopf B, Smola AJ, Williamson RC, Bartlett PL. New support vector algorithms. Neural Comput. 2000; 12: 1207–1245. https://doi.org/10.1162/089976600300015565.

15. Robin X, Turck N, Hainard A, Tiberti N, Lisacek F, Sanchez JC, et al. pROC: an open-source package for R and S+ to analyze and compare ROC curves. BMC bioinformatics. 2011; 12(1): 77. https://doi.org/10.1186/1471-2105-12-77 PMID:21414208.

16. Hall MW, Gavrilin MA, Knatz NL, Duncan MD, Fernandez SA, Wewers MD. Monocyte mRNA phenotype and adverse outcomes from pediatric multiple organ dysfunction syndrome. Pediatr Res. 2007; 62(5):597. https://doi.org/10.1203/PDR.0b013e3181559774 PMID: 17805202.

17. Boettcher S, Manz MG. Regulation of inflammation-and infection-driven hematopoiesis. Trends Immunol. 2017; 38(5): 345–57. https://doi.org/10.1016/j.it.201701.004 PMID: 28216309.

18. Meng C, Liu G, Mu H, Zhou M, Zhang S, Xu Y. Amphiregulin may be a new biomarker of classically activated macrophages. Biochem Biophys Res Commun. 2015; 466(3): 393–9. https://doi.org/10.1016/j.bbrc.2015.09.037 PMID: 26365345.

19. Zaiss DM, Gause WC, Osborne LC, Artis D. Emerging functions of amphiregulin in orchestrating immunity, inflammation, and tissue repair. Immunity. 2015; 42(2): 216–26. https://doi.org/10.1016/j.immuni.2015.01.020 PMID: 25692699.

20. Fujiu K, Shibata M, Nakayama Y, Ogata F, Matsumoto S, Noshita K, et al. A heart– brain–kidney network controls adaptation to cardiac stress through tissue macrophage activation. Nat Med. 2017; 23(5): 611. https://doi.org/10.1038/nm.4326 PMID: 28394333.

21. Campos EI, Reinberg D. Histones: annotating chromatin. Annu Rev Genet. 2009; 43: 559–599. https://doi.org/10.1146/annurev.genet.032608.103928 PMID: 19886812.

22. Grant PA. A tale of histone modifications. Genome Biol. 2001; 2(4): reviews0003–1. https://doi.org/10.1186/gb-2001-24-reviews0003 PMID: 11305943.

23. Jenuwein T, Allis CD. Translating the histone code. Science. 2001; 293(5532): 1074–80. https://doi.org/10.1126/science.1063127 PMID: 11498575.

24. Falvo JV, Jasenosky LD, Kruidenier L, Goldfeld AE. Epigenetic control of cytokine gene expression: regulation of the TNF/LT locus and T helper cell differentiation. Adv Immunol. 2013; 118: 37–128. https://doi.org/10.1016/B978-0-12-407708-9.00002-9 PMID: 23683942.

25. Ekaney ML, Otto GP, Sossdorf M, Sponholz C, Boehringer M, Loesche W, et al. Impact of plasma histones in human sepsis and their contribution to cellular injury and inflammation. Crit Care. 2014; 18(5): 543. https://doi.org/10.1186/s13054-014-0543-8 PMID: 25260379.

26. Alhamdi Y, Abrams ST, Cheng Z, Jing S, Su D, Liu Z, et al. Circulating histones are major mediators of cardiac injury in patients with sepsis. Crit Care Med. 2015; 43(10): 2094–103. https://doi.org/10.1097/CCM.0000000000001162 PMID: 26121070.

27. Wen Z, Jin Y, Jiang X, Sun M, Arman N, Wen T, et al. Extracellular histones indicate the prognosis in patients undergoing extracorporeal membrane oxygenation therapy. Perfusion. 2019; 34(3): 211–216. https://doi.org/10.1177/0267659118809557 PMID: 30370815.

28. Xu J, Zhang X, Pelayo R, Monestier M, Ammollo CT, Semeraro F, et al. Extracellular histones are major mediators of death in sepsis. Nat Med. 2009; 15(11): 1318. https://doi.org/10.1038/nm.2053 PMID: 19855397.

29. Czaikoski PG, Mota JM, Nascimento DC, Sônego F, Melo PH, Scortegagna GT, et al. Neutrophil extracellular traps induce organ damage during experimental and clinical sepsis. PloS one. 2016; 11(2): e0148142. https://doi.org/10.1371/journal.pone.0148142 PMID: 26849138.

30. Nakazawa D, Kumar SV, Marschner J, Desai J, Holderied A, Rath L, et al. Histones and neutrophil extracellular traps enhance tubular necrosis and remote organ injury in ischemic AKI. J Am Soc Nephrol. 2017; 28(6): 1753–68. https://doi.org/10.1681/ASN.2016080925 PMID: 28073931.

31. Li RH, Tablin F. A comparative review of neutrophil extracellular traps in sepsis. Front Vet Sci. 2018; 5. https://doi.org/10.3389/fvets.2018.00291 PMID: 30547040.

